# Does OMOP CDM Conversion Improve Cross-Country Comparability of Real-World Data? A Benchmark Study in Breast Cancer and Amyotrophic Lateral Sclerosis

**DOI:** 10.64898/2026.07.06.26357353

**Authors:** Mohamed Aborageh, Monika Roberta Korcinska Handest, Istvan Bakos, Blair Rajamaki, Célia Silva, Erzsébet Horváth-Puhó, Liisa Pylkkänen, Catarina Venda, Manuel Lentzen, Cornelia Becker, Joana Fernandes, Anne Paakinaho, Thuan Vo, Britta Haenisch, Sirpa Hartikainen, Anna-Maija Tolppanen, Claudia Furtado, Holger Fröhlich, Vera Ehrenstein

**Affiliations:** Fraunhofer Institute for Algorithms and Scientific Computing SCAI, Department of Biomedical AI & Data Science, Sankt Augustin, Germany; University of Bonn, Bonn-Aachen International Center for IT (b-it), Bonn, Germany; Danish Medicines Agency, Data and Data Analytic Center (DAD), Copenhagen, Denmark; Aarhus University and Aarhus University Hospital, Department of Clinical Epidemiology, Center for Population Medicine, Aarhus, Denmark; University of Eastern Finland, Faculty of Health Sciences, School of Pharmacy, Kuopio, Finland; INFARMED – National Authority of Medicines and Health Products, I.P., Lisbon, Portugal; University of Turku, Department of Oncology, Turku, Finland; Federal Institute for Drugs and Medical Devices, Research Division, Bonn, Germany; NOVA University Lisbon, NOVA National School of Public Health, Lisbon, Portugal; University of Bonn, University Hospital Bonn, Institute for Digital Medicine, Bonn, Germany; German Center for Neurodegenerative Diseases (DZNE), Bonn, Germany; University of Bonn, University Hospital Bonn, Center for Translational Medicine, Bonn, Germany

**Keywords:** Real-world data, Public health surveillance, Data harmonisation, Multi-country comparison, Health data governance, Rare diseases, Cancer epidemiology, European health systems, OMOP

## Abstract

**Background:** Real-world data (RWD) from different countries are increasingly used to support regulatory, health technology assessment (HTA), and population-level evidence generation. However, cross-country analyses are challenged by differences in data provenance, healthcare systems, coding practices, completeness, and clinical workflows. The Observational Medical Outcomes Partnership (OMOP) common data model (CDM) is widely used to harmonise heterogeneous RWD sources, but its ability to improve comparability of downstream epidemiological analyses relative to native source data across countries requires empirical evaluation.

**Methods:** We examined RWD from Denmark, Finland and Portugal in their ability to capture epidemiology of female breast cancer (BC) and amyotrophic lateral sclerosis (ALS), exemplifying, respectively, a common disease with established treatment modalities and high survival and a rare fatal disease with scarce treatment options. To enable head-to-head comparison on a semantic level, data were mapped to the OMOP CDM. Data in the native format were used for comparison. In a downstream analysis, we examined disease epidemiology, patient characteristics, treatment, and survival.

**Results:** OMOP conversion enabled a common analytical framework across countries and supported semantically aligned comparisons of key epidemiological and clinical variables. However, cross-country comparability was influenced by differences in data provenance, population coverage, coding practices, availability of clinical details, treatment capture, and healthcare-system-specific workflows. Iterative comparison with native data and external clinical evidence was necessary to identify mapping issues, assess information loss, and ensure high semantic fidelity of the converted data. Overall, OMOP-based estimates were highly consistent with native-data analyses and existing clinical expectations, but residual discrepancies reflected both source-data heterogeneity and decisions in the Extract, Transform, Load (ETL) workflow design.

**Conclusions:** OMOP CDM conversion facilitates semantically meaningful cross-country analyses of RWD by mapping heterogeneous source data to a common structure and standardised vocabularies. However, CDM conversion does not eliminate heterogeneity in the underlying data-generating processes and cannot substitute for study-specific data quality and fitness-for-purpose assessment. Robust use of harmonised RWD for regulatory, HTA, or population-level evidence generation requires iterative benchmarking against native data, clinical expertise, and data-science expertise to support valid interpretation across countries.

## Background

Randomised clinical trials (RCTs) remain the gold standard and the regulatory requirement for establishing the safety and efficacy of medications prior to authorisation. However, their strict eligibility criteria, limited sample sizes, and highly controlled settings aimed to maximise internal validity constrain external validity of the findings in routine clinical practice. Real-world evidence (RWE) stemming from analysing routinely collected healthcare data, when used judiciously, is potentially a valuable complement to RCTs in regulatory decision-making and health technology assessment (HTA) [1]. While the use of real-world data (RWD) is well established in pharmacovigilance, its application earlier in the product lifecycle – particularly to support pre-authorisation decision-making – remains an evolving area, though increasingly recognised for selected purposes by the European Medicines Agency [1–5].

Sources of secondary RWD from different countries vary in data provenance and flow, coding practices, completeness, and coverage. These affect comparability, interpretability, and regulatory utility of RWD [6, 7]. The Real4Reg project is a European initiative that explores, through representative use cases, how RWD supported by artificial intelligence (AI) may inform regulatory and HTA decision-making in different European countries. One Real4Reg use case assessed the feasibility of using European RWD sources in the pre-authorisation and evaluation phase of treatments in regulatory practice [8]. The use case focuses on epidemiology and clinical course of two conditions with contrasting epidemiological and clinical characteristics: female breast cancer (BC), the most common malignancy in women [9, 10], and amyotrophic lateral sclerosis (ALS), a rare neurodegenerative disease [11, 12].

The Observational Medical Outcomes Partnership (OMOP) common data model (CDM), originally developed in the United States, has been increasingly adopted in Europe for semantically harmonising RWD, also in regulatory settings [1, 13–17]. However, its contribution to cross-country comparability is often assumed rather than empirically evaluated. Existing work has demonstrated the feasibility of OMOP mapping, assessed information loss during conversion, established data quality assessment workflows, and shown that OMOP enables large-scale distributed analyses [18]. However, comparatively little evidence exists on whether transformation to OMOP improves the comparability of downstream epidemiological estimates across countries relative to analyses conducted on native source data. This distinction is important because CDM conversion can harmonise data structure and terminology, but cannot eliminate differences in clinical practice, healthcare systems, coding incentives, data completeness, or source-specific data provenance.

In this study, we benchmarked native and OMOP-based analyses across three European registry systems in two contrasting disease areas, female BC and ALS. Our first aim was to document the harmonisation of diverse European RWD sources to the OMOP CDM and assess the quality of the mapped data. Our second aim was to evaluate whether OMOP conversion improves cross-country comparability of downstream descriptive analyses, using native-data analyses as the benchmark. By comparing reproducibility of estimates before and after harmonisation, we aimed to identify both the benefits of CDM-based semantic harmonisation and the residual discrepancies introduced or preserved by Extract, Transform, Load (ETL) decisions, vocabulary mappings, and country-specific data structures.

## Methods

### Study Design and Data Sources

We conducted a multinational cohort study. The data used in this study came from national healthcare and administrative population registers and databases in three European countries: Denmark, Finland and Portugal [19–21]. Detailed overview of each data source is provided in Supplementary Table 1.

### Study Population and Variables

Patients with BC and ALS were identified within country-specific study periods according to data recency at the time of extraction (Denmark and Finland: 2000–2021; Portugal: 2005–2022). Eligible patients were adults (age ≥18 years) with a first-recorded diagnosis of BC or ALS during the study period. The date of the diagnosis was the index date. Exclusion criteria, applied during the 5 years before the index date, were prevalent condition and lack of continuous country residence. For BC, an additional exclusion criterion was a history of any other malignancy, except carcinoma in situ of the cervix, basal cell carcinoma of the skin, or squamous cell carcinoma of the skin.

Baseline variables varied by country and included age, sex, socioeconomic status, comorbidities, and medication use in the 5 years before the index date. Disease-specific characteristics (e.g., cancer receptor status, tumour stage, metastases, or ALS-related treatments) and indicators of disease progression (e.g., survival, hospitalisations, intensive care admissions, and respiratory failure) were ascertained during the clinically relevant period for progression, defined as the period after the index date and up to the end of follow-up.

### Data Harmonisation Workflow

To benchmark the CDM-transformed results, data in each country were analysed in native format using R scripts. Native data from each country underwent data management and curation procedures, including country-specific quality checks, coding of study variables, and preparation of analysis-ready datasets. Subsequently, country-specific data were transformed into the OMOP CDM version 5.4 using a structured ETL workflow [22]. This transformation harmonised coding systems, database structures, and national registry conventions across datasets, enabling standardised analyses within a downstream common analytical framework (syntactic and semantic harmonisation). The transformation workflow is shown in Figure 1.

**Figure 1:**
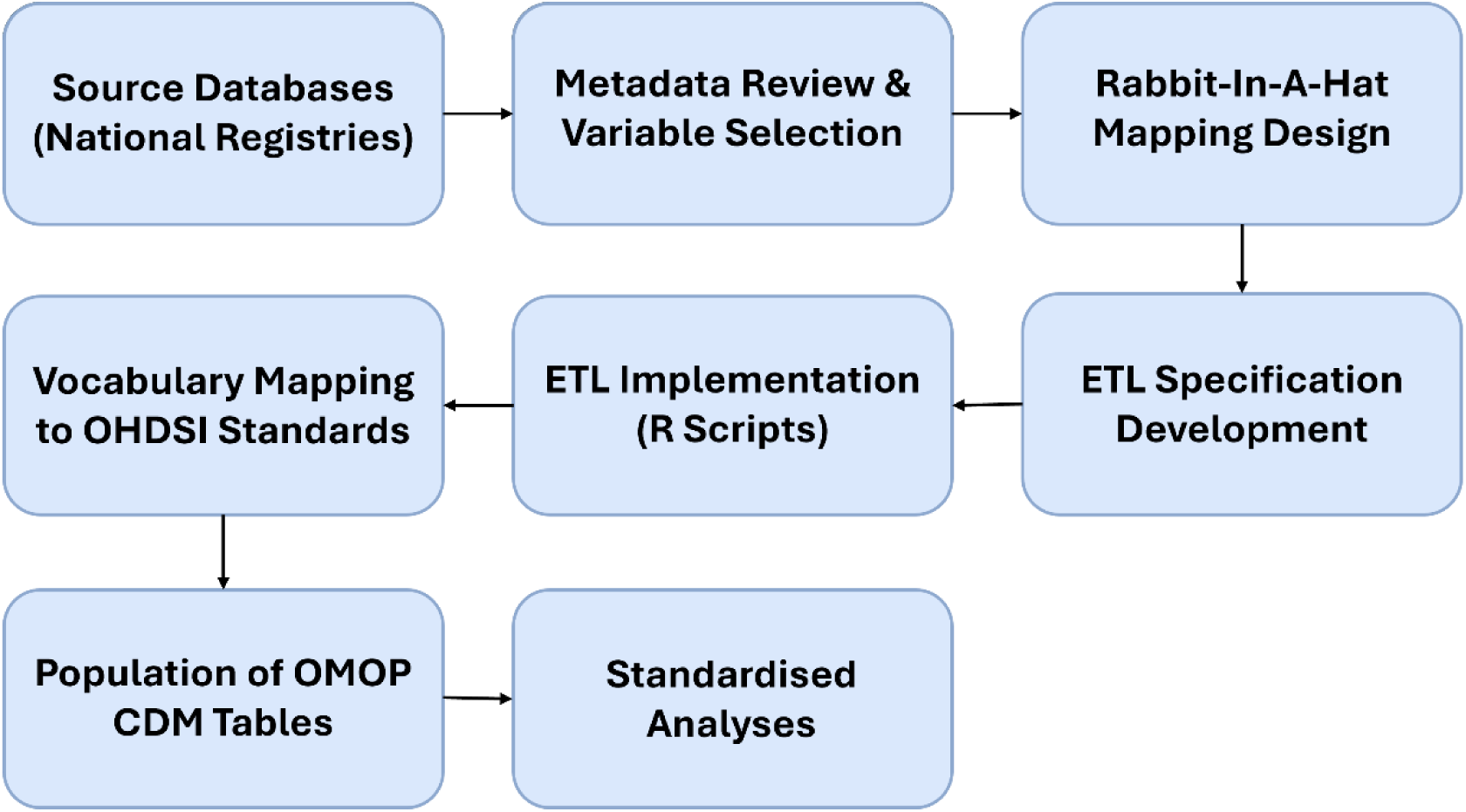
Overview of the Extract–Transform–Load (ETL) process used to harmonise country-specific data into the OMOP Common Data Model (CDM).

The OMOP CDM structures healthcare information within a relational database composed of standardised tables organised into six main categories:

1. Clinical data tables, which record demographics and patient-level healthcare events such as deaths (including date and cause), healthcare visits with associated diagnoses or conditions, drug prescriptions or dispensing, use of medical devices, and surgical or medical procedures
2. Health economics tables, containing financial information linked to clinical events, insurance details, and derived continuous exposure or condition periods
3. Standardised vocabulary tables, which ensure semantic consistency by defining core vocabularies, concept relationships, hierarchies, and usage types
4. Metadata tables, documenting data transformation processes
5. Health system tables, which describe locations, care sites, and providers
6. Derived elements, including cohorts as well as aggregated drug, condition, and dose era records.

The following subset of the OMOP tables was necessary to address the aims of this study: person, death, visit_occurrence, condition_occurrence, drug_exposure, procedure_occurrence, measurement, observation_period, concept, concept_relationship and vocabulary (Supplementary Material 1).

The OMOP CDM supports the use of standardised vocabulary – such as SNOMED CT and RxNorm/RxNorm Extension – to represent clinical concepts in a harmonised manner. Accordingly, concepts from the source datasets were first mapped to the appropriate standardised terms prior to data transformation. In Denmark, clinical coding is standardised through the Danish Healthcare Classification System (Sundhedsvæsenets Klassifikationssystem, SKS), which covers the Danish version of the International Classification of Diseases, 10^th^ Revision (ICD-10) classification for diagnoses and symptoms and the NOMESCO Classification of Surgical Procedures (NCSP) for surgical procedures [23]. The Anatomical Therapeutic Chemical (ATC) classification was used for the classification of drugs. For Finland, vocabulary files developed within the FinOMOP and FinnGen initiatives [24, 25] were utilised; these extend the OMOP Athena vocabularies and incorporate certain national ICD codes not covered by ICD-10 or ICD-10-CM and special reimbursement entitlement codes (erityiskorvausoikeus, ERKO), which are not fully represented in the standard vocabularies. In Portugal, most registries employ vocabularies already included in Athena, such as ATC, ICD-9-CM and ICD-10-CM, minimising the need for external mapping resources (Supplementary Material 2). In Denmark, the tumour size, lymph nodes, metastasis (TNM) breast cancer stage components recorded separately in the native system were combined during transformation to derive a unified stage. In Finland, stage information was available through a local classification rather than as a directly comparable TNM-derived stage variable. In Portugal, cancer stage (0–IV) was based on the TNM staging system.

The data harmonisation process began with a systematic review of the available metadata and data dictionaries provided by each data partner. Based on this information, all variables required for the study were identified and mapped from source tables to the corresponding OMOP CDM domains. Initial source-to-CDM table mappings were defined using the Rabbit-In-A-Hat tool provided by OHDSI [15], allowing data custodians to review and confirm the proposed mappings prior to ETL implementation.

Following confirmation of the mapping structure, an ETL specification was developed describing the transformation rules required to convert the source data to the OMOP CDM format. This specification included definitions of source-to-CDM table mappings as well as mappings of local coding systems to standardised OHDSI vocabularies. ETL scripts were implemented in R designed to facilitate reuse across datasets from different countries. This modular approach allowed the same transformation logic to be applied across all data sources while accommodating minor adjustments required to address structural differences between national registries and variations in local coding systems. Subsequently, local clinical codes were mapped to the OMOP standardised vocabularies (SNOMED, ICD-10, ICD-9 and ATC) to ensure semantic harmonisation between datasets. Finally, the transformed data were loaded into OMOP CDM tables, creating standardised databases for shared analysis across participating countries.

The transformation process followed an iterative Extract-Transform-Load (ETL) workflow tailored to the structure and metadata of each participating dataset. An initial ETL specification was developed based on the metadata and data dictionaries provided by each data partner, defining table mappings, variable transformations, and vocabulary alignment strategies prior to direct data access. ETL scripts were implemented using modular R functions designed to facilitate reuse across countries while accommodating dataset-specific adaptations. This modular approach allowed consistent handling of common OMOP domains (e.g., person, condition, drug exposure, visit occurrence), while enabling targeted customisation where national coding systems or structural differences required deviations from the initial specification.

The ETL logic was refined to reflect the structure, completeness, and granularity of the source data, including adjustments to date handling, identifier linkage, and concept-mapping rules. These refinements were informed by exploratory analyses and iterative benchmarking against the native-format outputs to ensure alignment between source and OMOP-derived representations.

Following transformation, the resulting OMOP datasets were systematically evaluated using the OHDSI Data Quality Dashboard, which assesses conformance, completeness, and plausibility across predefined CDM checks. Identified issues were reviewed in collaboration with the data partners, and ETL procedures were updated where necessary to address structural inconsistencies or mapping gaps prior to final analysis.

The quality of the OMOP-transformed datasets was evaluated using the OHDSI Data Quality Dashboard separately for the BC and ALS subsets. The dashboard evaluates multiple dimensions of data quality, including conformance, completeness, and plausibility. Conformance checks assess whether the transformed data adhere to the structural requirements of the OMOP Common Data Model (e.g., correct table structures, required fields, and relational integrity). Completeness checks evaluate whether expected data elements are populated and whether source codes have been successfully mapped to standard concepts. Plausibility checks assess whether recorded values and temporal relationships are clinically and logically consistent.

### Statistical Analysis

The epidemiology of disease was described with age- and sex-stratified incidence rates. Incidence rate was computed as the number of new cases per 100,000 person-years. Rates were standardised by age and sex using the 2013 European Standard Population (EU-27) and reported with 95% confidence intervals [26]. The baseline characteristics of BC and ALS populations were described using descriptive statistics (means, medians, frequencies, percent). Both all-cause and disease-specific mortality was assessed. The clinical course was described using prevalence. Survival probabilities were estimated with the Kaplan–Meier method and plotted with 95% confidence intervals.

Analyses conducted on the native datasets were implemented in R using standard packages with standardised scripts applied locally in each country to calculate incidence, prevalence and survival [27].

Analyses performed in OMOP-CDM–transformed datasets were benchmarked against analyses on curated data in native format. Details of the operational definitions, analytical algorithms, statistical packages, and quality control procedures are available in Supplementary Material 3.

### Quality control

All analytic and visualisation scripts were developed in R and independently validated. Independent validation was performed by a second data scientist who reviewed the code, replicated the analyses, and verified concordance of the results. Scripts were developed in adherence to the analysis data model (ADaM) structure to ensure reproducibility, traceability, and consistent execution across country-specific data nodes [28]. Country teams performed local programming quality control, and results were benchmarked against published evidence and clinical expertise. The full study underwent multiple rounds of review by the research consortium.

### Ethical Approvals and Data Protection

In Denmark and Finland the ethics approval or informed consent were not required as the study was conducted under the legislation on secondary use of health and social data. Research team had access to pseudonymised data and study participants were not contacted All analyses were conducted within secure national data environments (Danish Health Data Authority, National Genome Center in Denmark and Findata in Finland. The Portuguese Ethics Committee for Clinical Research (CEIC) approved the study plan (Deliberation no. 2024-RP-04-13, 12 April 2024). All data were pseudonymised before access by the research team, and all analyses were conducted within secure national data environments at INFARMED, I.P. Access, in all countries was limited to authorised researchers in accordance with national regulations. Only aggregated results were reported, following national disclosure rules to prevent re-identification of individuals and privacy breaches. Minimum cell-count thresholds were applied (Finland: <3; Denmark: *<*5; Portugal: *<*10;), with Denmark additionally rounding person counts to the nearest five in the native analysis results. All derived descriptive statistics, incidence and prevalence measures, and mortality estimates were calculated from unrounded source data.

## Results

### OMOP Data Quality Assessment

Across 2,201 executed checks per subset, overall pass rates for plausibility, conformance and completeness were 93-97% for the BC dataset and 94-97% for the ALS dataset. Tables 1 & 2 present the results generated by the assessment tool for both the ALS and BC subsets in all three countries. More detailed results for the quality checks can be found in Supplementary Tables 4-9.

**Table 1:** Data quality assessment of OMOP-transformed ALS subset from the registries data per country.

|  | Verification |  |  | Validation |  |  | Total |  |  |
| --- | --- | --- | --- | --- | --- | --- | --- | --- | --- |
|  | Denmark | Finland | Portugal | Denmark | Finland | Portugal | Denmark | Finland | Portugal |
| Plausibility | 507 (98%) | 505 (98%) | 513 (100%) | 291 (100%) | 288 (99%) | 277 (95%) | 798 (99%) | 793 (98%) | 790 (98%) |
| Conformance | 747 (93%) | 747 (93%) | 784 (97%) | 116 (97%) | 119 (100%) | 118 (99%) | 863 (93%) | 868 (94%) | 902 (97%) |
| Completeness | 402 (89%) | 405 (90%) | 440 (98%) | 18 (95%) | 12 (63%) | 14 (74%) | 420 (90%) | 417 (89%) | 454 (97%) |
| **Total** | 1656 (93%) | 1657 (94%) | 1737 (98%) | 425 (99%) | 419 (98%) | 409 (95%) | 2081 (95%) | 2076 (94%) | 2146 (97%) |

**Table 2:** Data quality assessment of OMOP-transformed BC subset from the registries data per country.

|  | Verification |  |  | Validation |  |  | Total |  |  |
| --- | --- | --- | --- | --- | --- | --- | --- | --- | --- |
|  | Denmark | Finland | Portugal | Denmark | Finland | Portugal | Denmark | Finland | Portugal |
| Plausibility | 507 (98%) | 505 (98%) | 515 (100%) | 288 (99%) | 277 (95%) | 281 (97%) | 795 (99%) | 782 (97%) | 796 (99%) |
| Conformance | 744 (92%) | 741 (92%) | 776 (96%) | 115 (97%) | 118 (99%) | 118 (99%) | 859 (93%) | 860 (93%) | 894 (96%) |
| Completeness | 401 (89%) | 403 (90%) | 449 (98%) | 18 (95%) | 15 (79%) | 14 (74%) | 419 (89%) | 414 (88%) | 457 (98%) |
| **Total** | 1652 (93%) | 1649 (93%) | 1776 (98%) | 421 (98%) | 414 (97%) | 409 (95%) | 2073 (94%) | 2056 (93%) | 2147 (97%) |

For both diseases, plausibility checks demonstrated 97-99% of checks passing overall, indicating high internal consistency of clinical values and temporal relationships. Conformance checks achieved pass rates of 93-97% in both subsets, reflecting good adherence to CDM structural specifications and relational integrity constraints. Completeness checks yielded pass rates of 88-98% for BC and 89-90% for ALS, with failures primarily attributable to partially populated optional fields or domains not uniformly captured in source data. Verification and validation checks showed consistent patterns across disease subsets, with validation checks achieving higher pass rates (BC: 95-98%; ALS: 95-99%) than verification checks (both 93-98%), suggesting stable structural implementation with limited residual inconsistencies.

No critical failures affecting primary key integrity, person-level linkage, or essential analytical domains were identified. Overall, these findings indicate that both OMOP-transformed subsets achieved high structural and semantic quality, supporting their suitability for cross-country descriptive analyses.

### Data Heterogeneity

Substantial diversity in data completeness, granularity, and recording practices was observed across the three countries and between the two conditions studied. For BC, incidence estimates in Denmark and Finland aligned well with published evidence, and demographic and comorbidity profiles were comparable [29, 30]. However, completeness of tumour stage and receptor-status data varied widely, with 20–80% missing stage information and limited availability of molecular characteristics outside of Denmark. For Portugal, incidence rates of breast cancer could not be reliably estimated due to missing linkage between the oncology registry and hospital records. Patients with breast cancer were identified using hospital records, while the oncology registry provided additional information; however, the registry included a larger population—partly due to the inclusion of patients treated in private hospitals—that could not be linked to other datasets (e.g., the National User Register) following anonymisation, thereby precluding integrated analysis.

For ALS, demographic and comorbidity distributions were broadly comparable across the three countries, although some differences may reflect variation in data capture across countries. In particular, cardiovascular disease was recorded less frequently in Denmark, which may partly relate to differences in the underlying data sources. Recording riluzole ALS treatment use was captured in Denmark and Finland with variable prevalence. Riluzole treatment was not captured in Portugal, since it is administered in hospitals, and hospital dispensings do not generate patient-level records. The granularity of diagnostic coding also differed, with ALS-specific diagnosis codes available in ICD-10 modifications used in Portugal and Denmark, but not in Finland, where operational definition of ALS therefore necessarily included other motor-neuron diseases.

Across all settings, RWD was linkable at the individual level through personal identifiers and were collected within universal healthcare systems. Data availability differed by source, purpose, and national policies. Some elements (age, sex, diagnoses deaths) were consistently available, while others - such as socioeconomic indicators or molecular tumour markers - were country-specific or restricted due to data-governance constraints (e.g., socioeconomic data are available in principle in Denmark, but were not analysed in the scope of this project). Data completeness varied by data type: cancer diagnoses recorded in mandatory national cancer registries with mandatory reporting were aligned with existing evidence. By contrast completeness of other variables by country (e.g., for breast cancer tumour stage, and ALS subtype or treatment).

### Comparison of Native vs OMOP Mapped Data

Overall, descriptive estimates were highly aligned across the two formats, with only minor differences. Minor discrepancies identified in early comparisons were resolved through verification of source-to-OMOP mappings and transformation logic prior to finalizing the OMOP datasets. Small percentage differences between native and OMOP-derived estimates remained, which could be expected in CDM transformations due to differences in coding systems, vocabulary mappings, and data standardisation procedures. Differences could also arise from one-to-many mapping relationships (e.g., ATC to multiple RxNorm concepts), which can increase the number of standardised drug records compared with native representations.

The study populations were comparable between native and OMOP-transformed datasets within each country, with no material differences observed in core demographic or comorbidity distributions. Table 3 presents the demographic and clinical characteristics of women with incident BC in the three countries. Age distributions were broadly consistent, with approximately 40% patients aged ≥65 years at diagnosis. Median age at diagnosis ranged from 60 years in Portugal to 64 years in Denmark. Psychiatric disorders were uncommon across all datasets (≤4%), whereas cardiovascular disease was more prevalent, ranging from 8% in Denmark and Portugal to 26% in Finland. Dementia was recorded in 1–3% of patients.

**Table 3:**
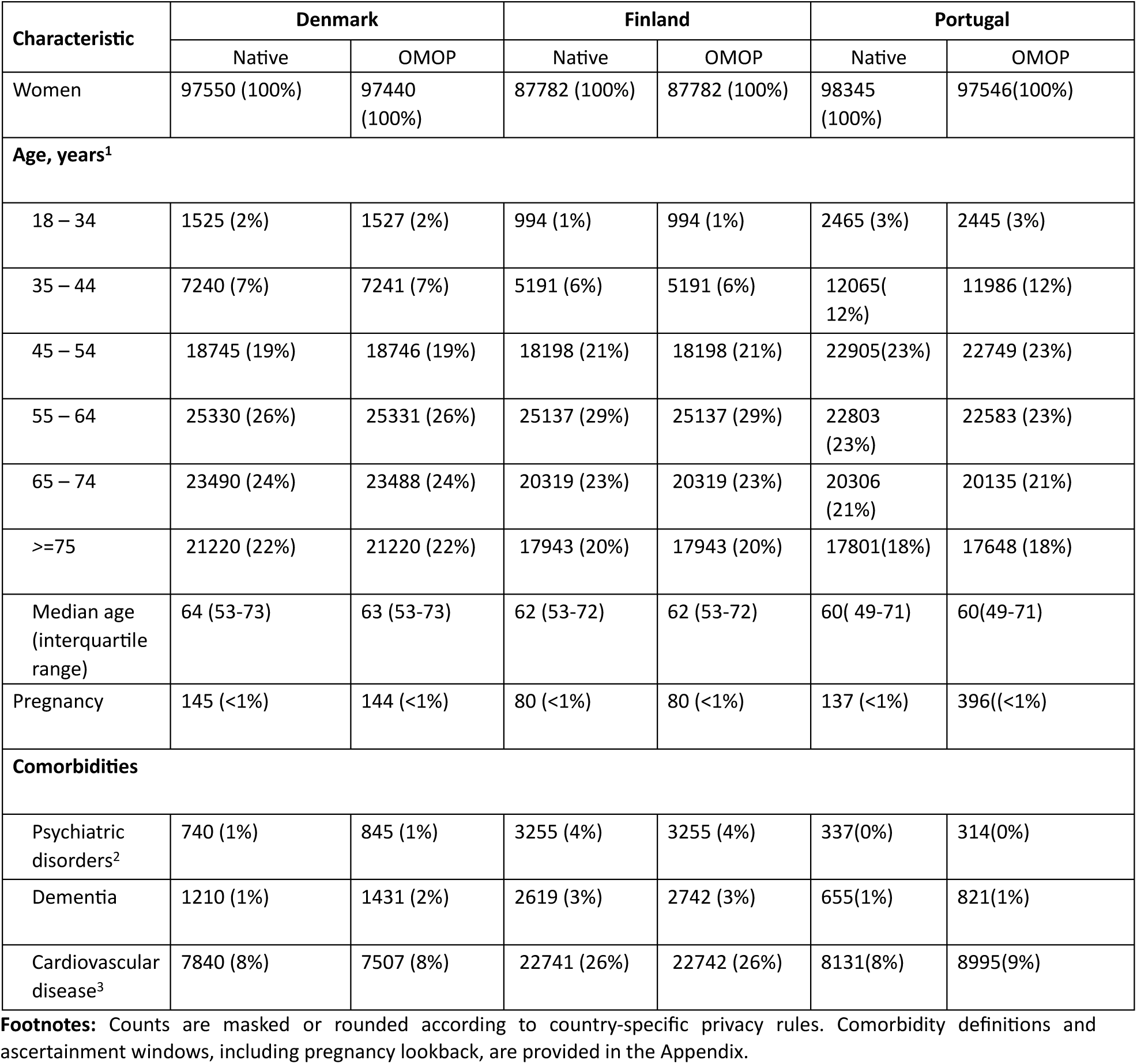
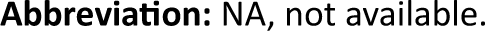
Selected characteristics of women with incident breast cancer in participating countries (Denmark and Finland, 2000–2021; Portugal, 2005–2022)

Additional demographic data, clinical characteristics, disease characteristics, and indicators of treatment and disease progression for BC can be found in supplementary Table 2.

Figure 2 represents breast cancer incidence rates over time in Denmark and Finland, showing broadly stable trends with small year-to-year changes and close alignment between native and OMOP estimates.

**Figure 2:**
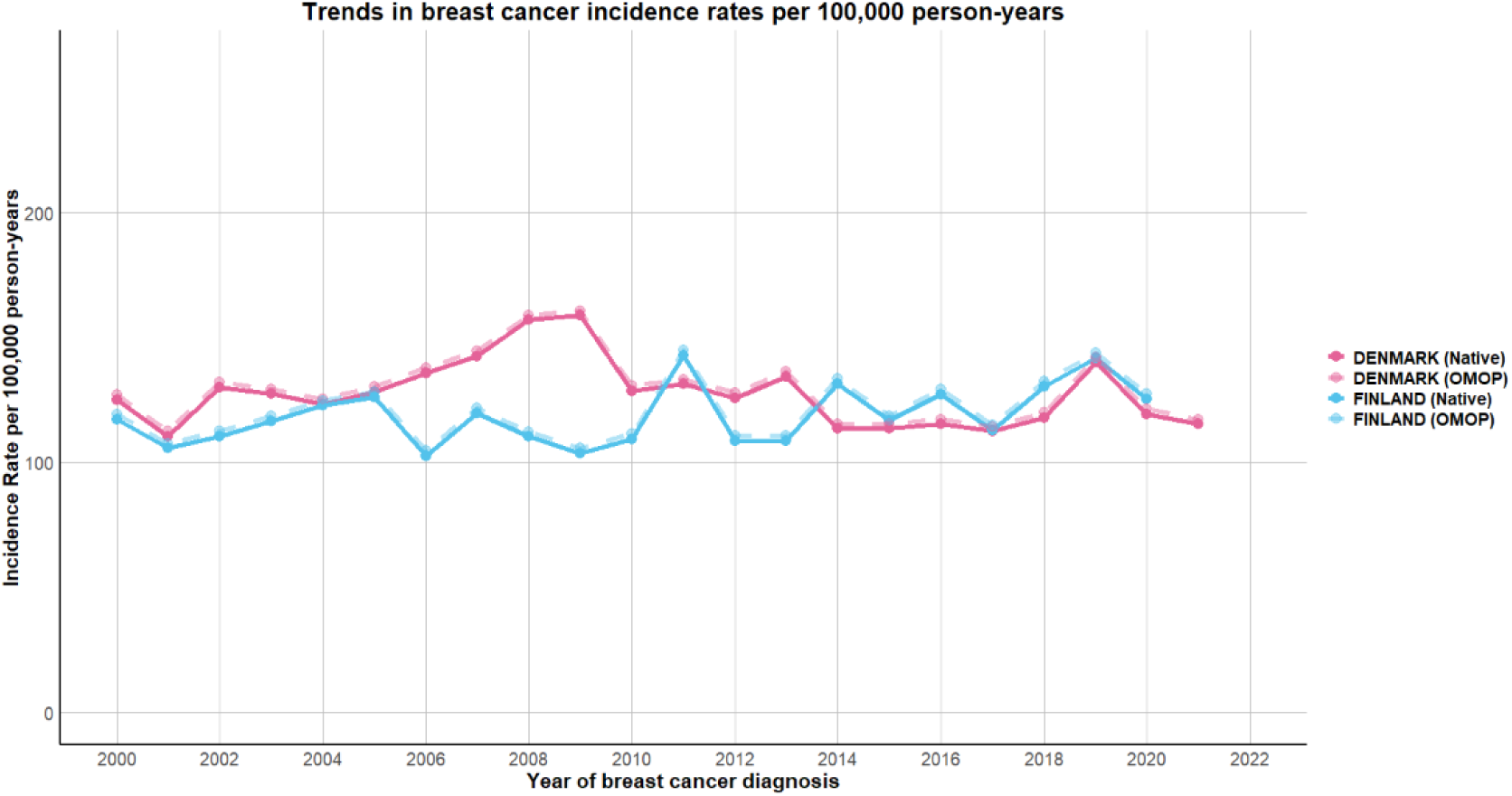
Annual age-standardized incidence rates of BC among men and women combined in native and OMOP data formats in Denmark and Finland, 2000–2021.

Figures 3 and 4 presents Kaplan–Meier survival curves for BC patients in Denmark and Finland during 2000-2021 period based on native data and OMOP-transformed data. The overall survival patterns are highly comparable between the two approaches, showing a gradual decline in survival over time with similar curve shapes. This indicates strong consistency between native and OMOP-based analyses for survival estimation.

**Figure 3:**
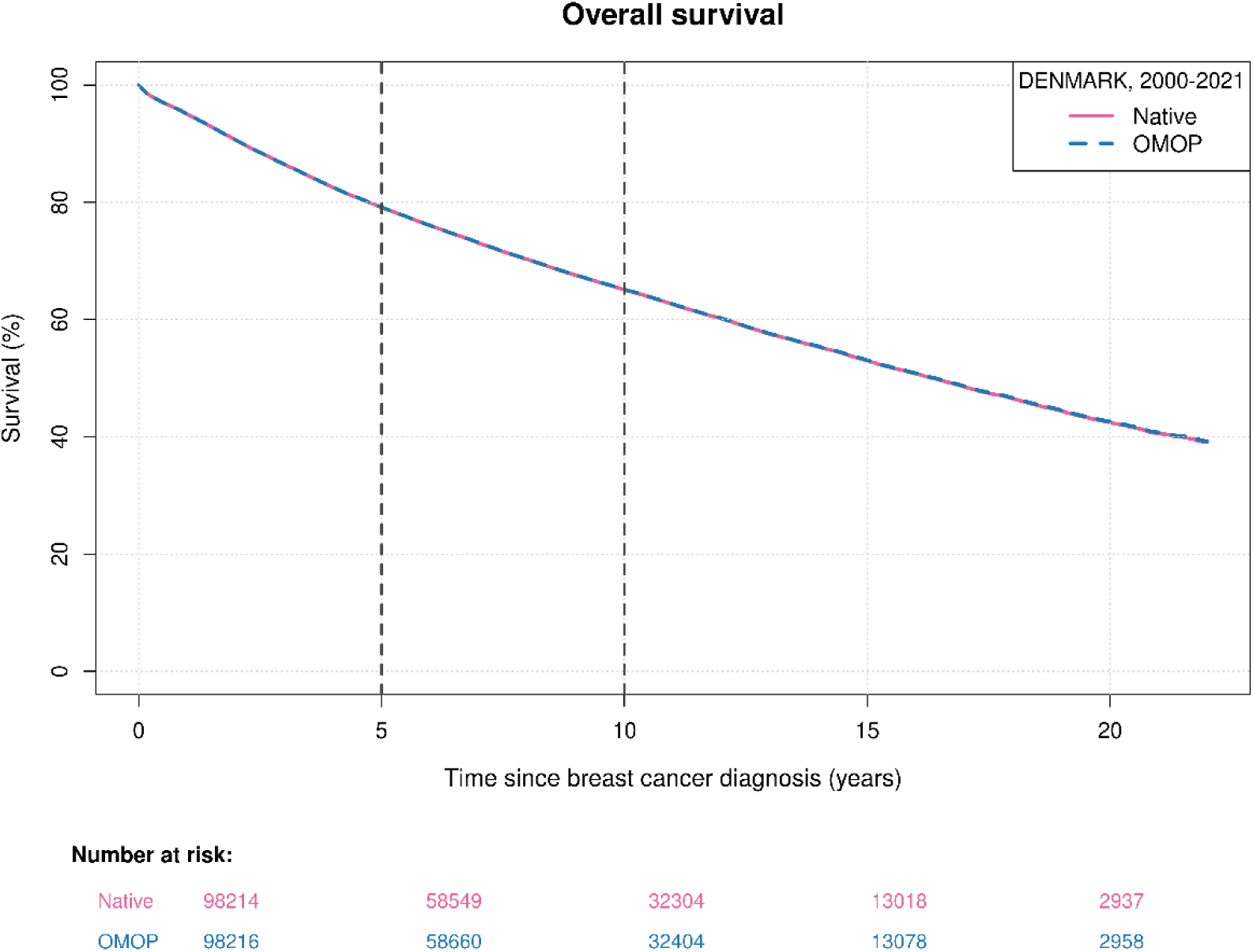
Kaplan-Meier survival curve of BC in the Danish native and OMOP data.

**Figure 4:**
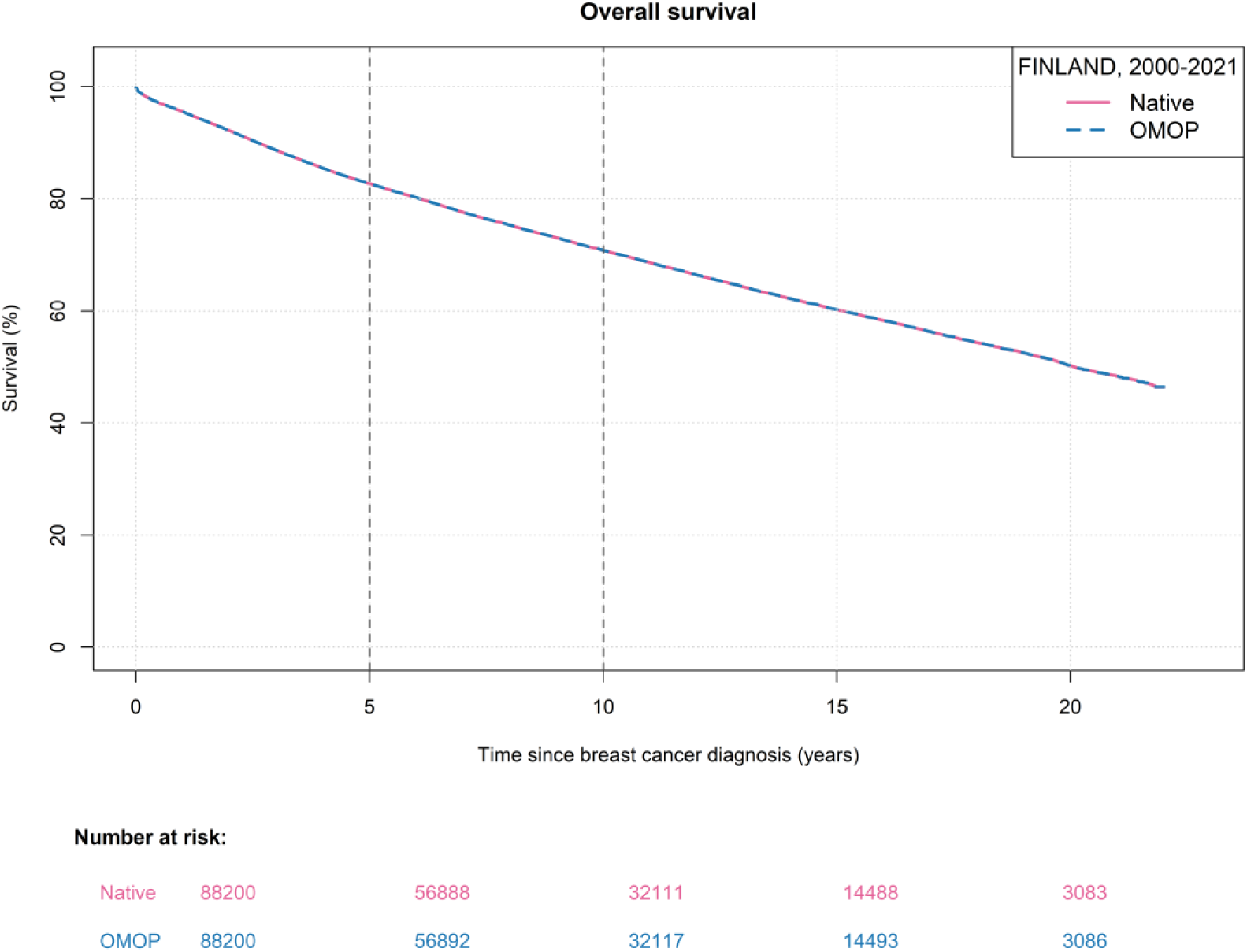
Kaplan-Meier survival curve of BC in the Finnish native and OMOP data.

Table 4 represents the demographic and clinical characteristics of ALS in the three countries. Sex distribution was comparable across countries, with 45–48% of patients being female. Most patients were aged ≥ 55 years at diagnosis, and median age ranged from 67 to 69 years across datasets. Prevalence of psychiatric disorders was 1–3%, and of dementia 4–6%. Recorded cardiovascular disease prevalence was 36% in Finland, 32% in Portugal, and 16% in Denmark. Regarding treatment patterns, riluzole was recorded in 58% of Danish and 30% of Finnish patients. In Portugal, individual-level data on riluzole use were unavailable because riluzole is primarily dispensed through hospital pharmacies and could not be captured at the patient level. Therefore, riluzole use observed in Denmark and Finland should be interpreted considering country-specific differences in ALS diagnostic coding, clinical practice, subtype-specific prescribing patterns, and medication data capture. In particular, the Finnish data included only outpatient pharmacy dispensings, whereas corresponding data were unavailable for Portugal. Rates of non-invasive ventilation and intensive care unit admission with mechanical ventilation were comparable between native and OMOP-based estimates in Denmark and Portugal; however, these data were not available for Finland.

**Table 4:**
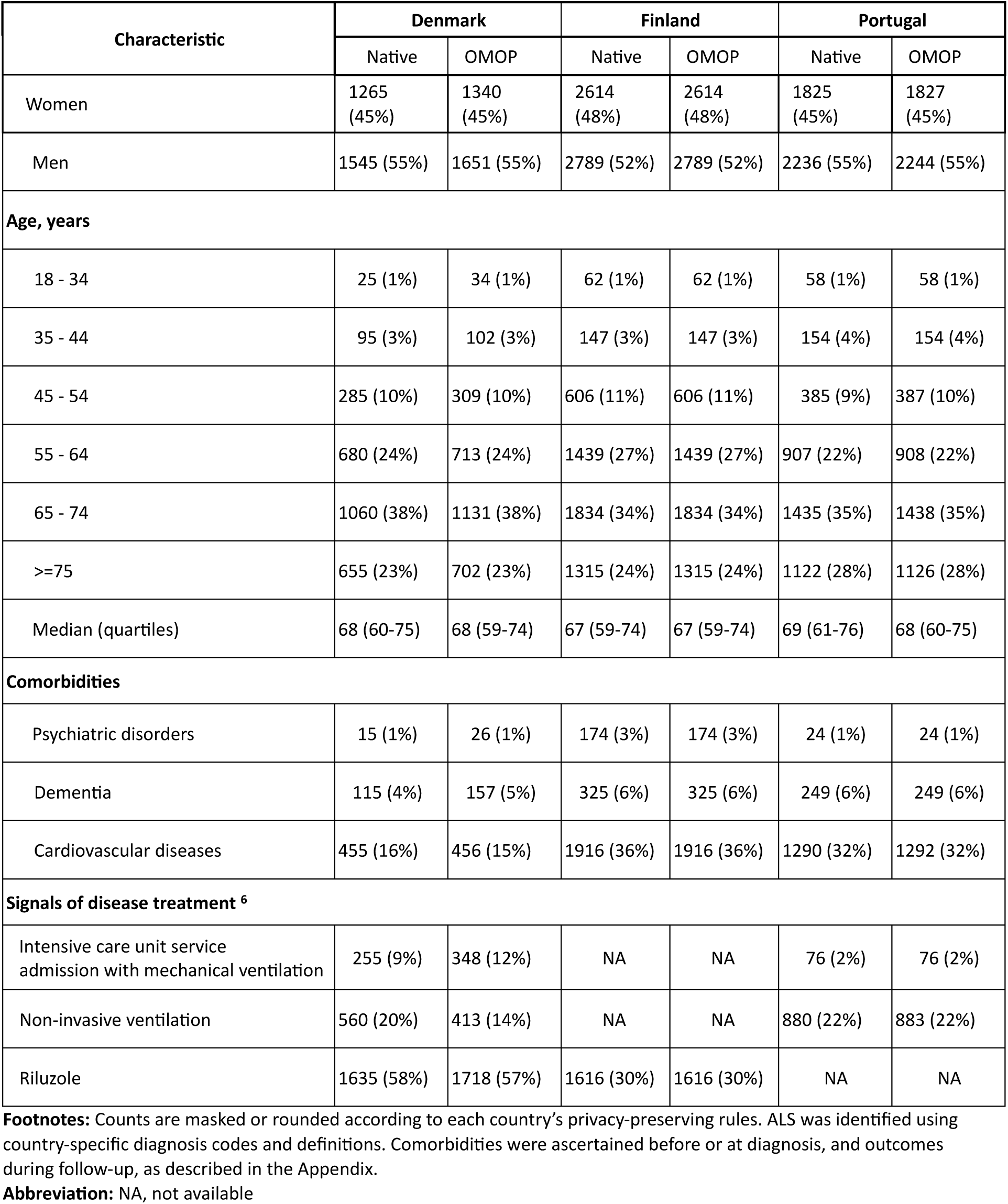
Selected characteristics of patients with incident ALS in participating countries (Denmark and Finland, 2000–2021; Portugal, 2005–2022)

Additional demographic data, clinical characteristics, disease characteristics, and indicators of treatment and disease progression for ALS can be found in supplementary Table 3.

Figure 5 represents ALS incidence rates over time in Denmark, Finland, and Portugal, showing higher rates in Finland, intermediate rates in Denmark, and lower rates in Portugal, with close alignment between native and OMOP estimates. The higher incidence observed in Finland may be partly explained by differences in case definitions, as ALS diagnoses in Denmark and Portugal were based only on ALS subtypes, whereas Finnish ALS cases included both subtypes and unspecified ALS cases

**Figure 5:**
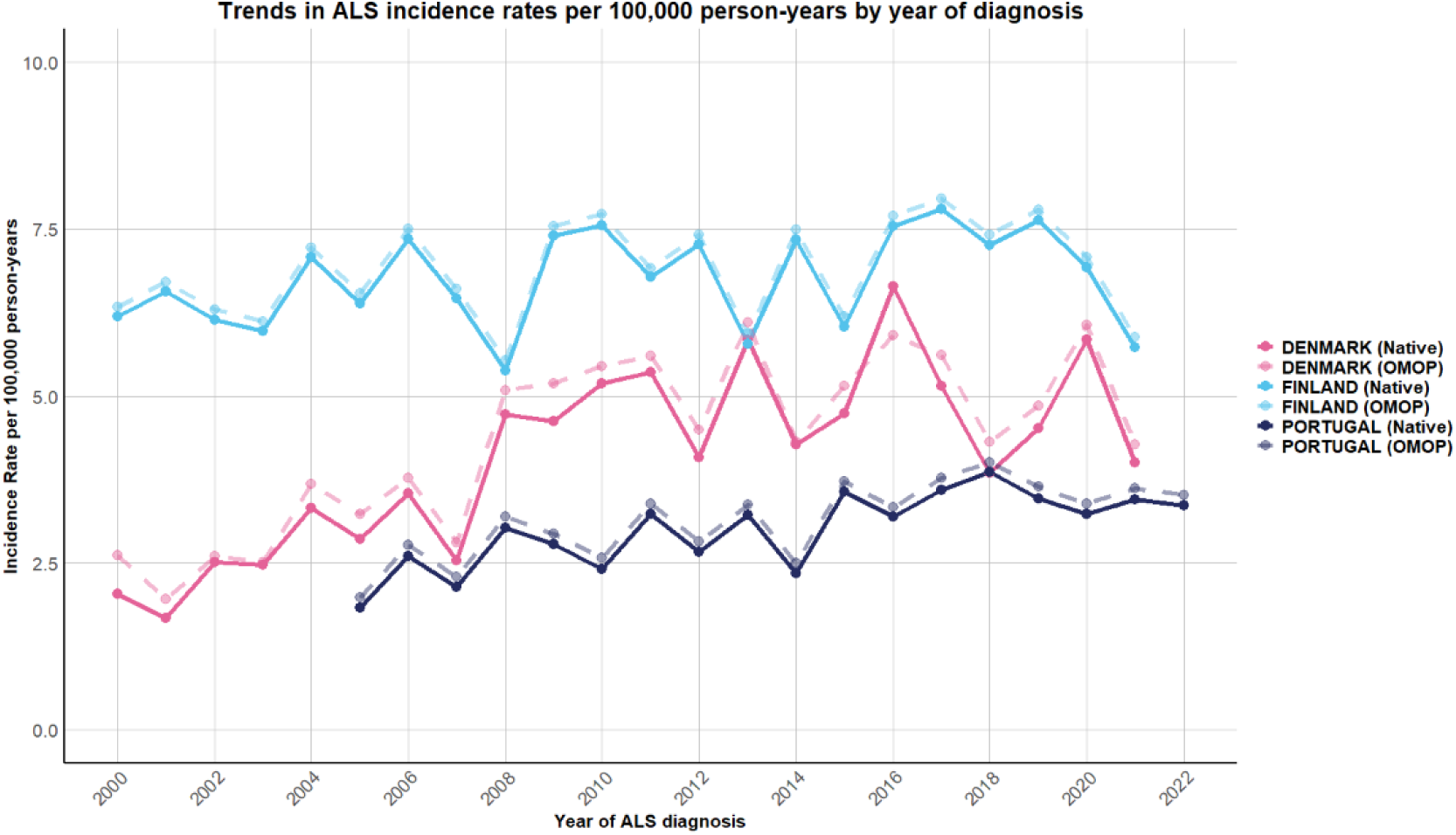
Annual incidence rates of ALS for Native and OMOP Formats in Denmark, Finland, and Portugal, 2000–2022.

Figures 6–8 illustrates Kaplan–Meier survival curves for ALS patients in all countries during the study period based on (a) native data and (b) OMOP-transformed data. The overall survival patterns are highly comparable between the two approaches, with both curves showing a rapid decline in survival during the early follow-up period.

**Figure 6:**
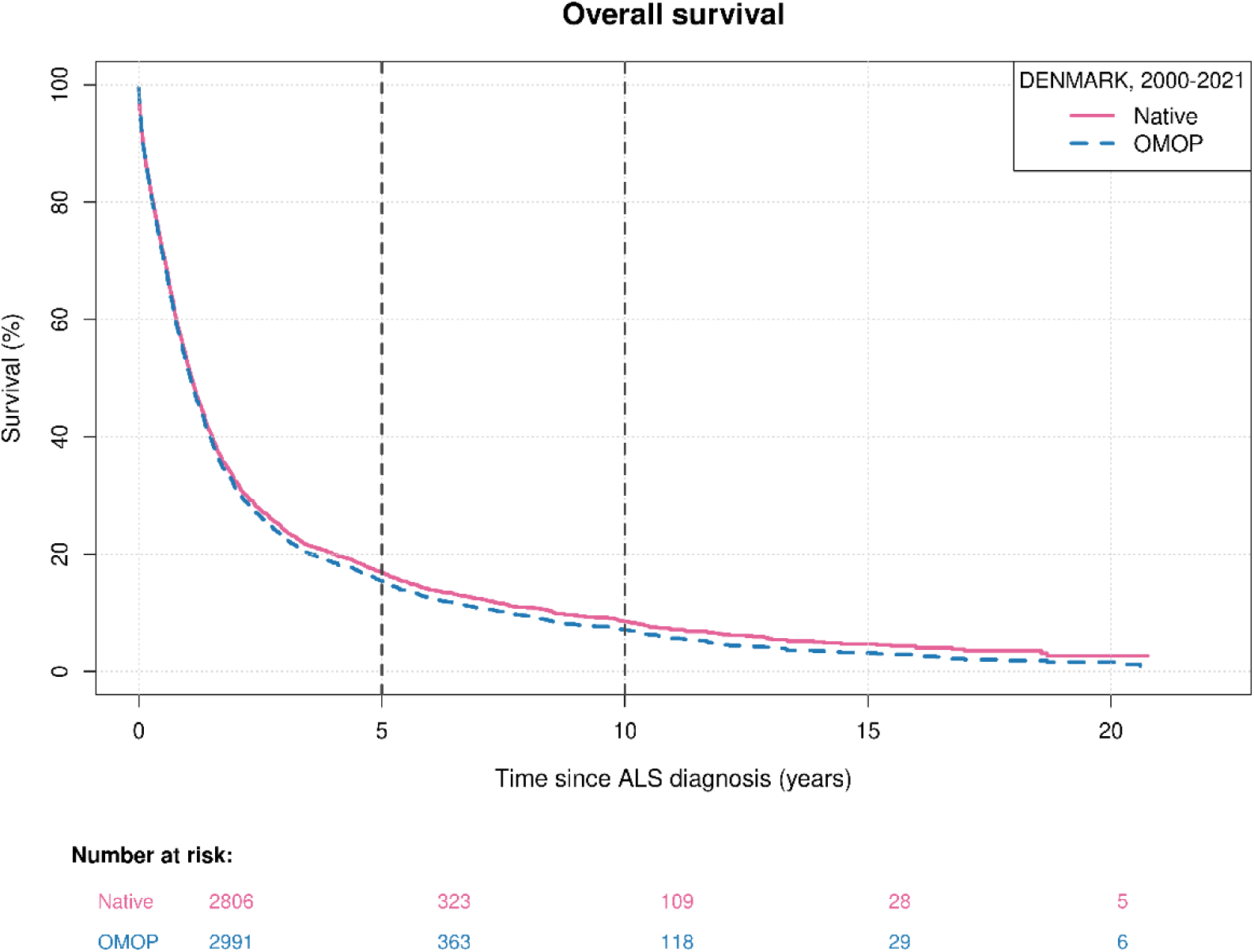
Kaplan-Meier survival curve of ALS in the Danish native and OMOP data.

**Figure 7:**
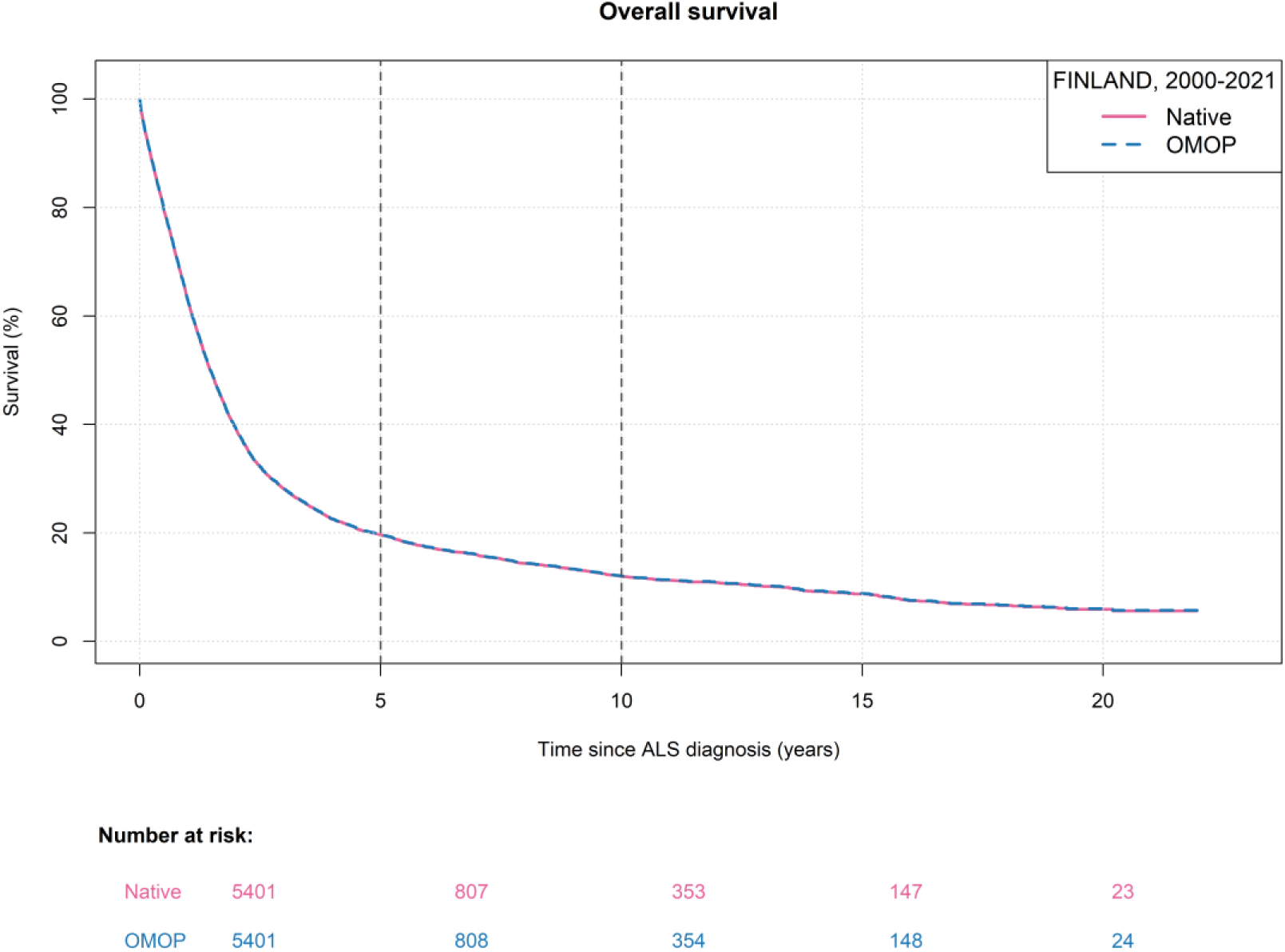
Kaplan-Meier survival curve of ALS in the Finnish native and OMOP data.

**Figure 8:**
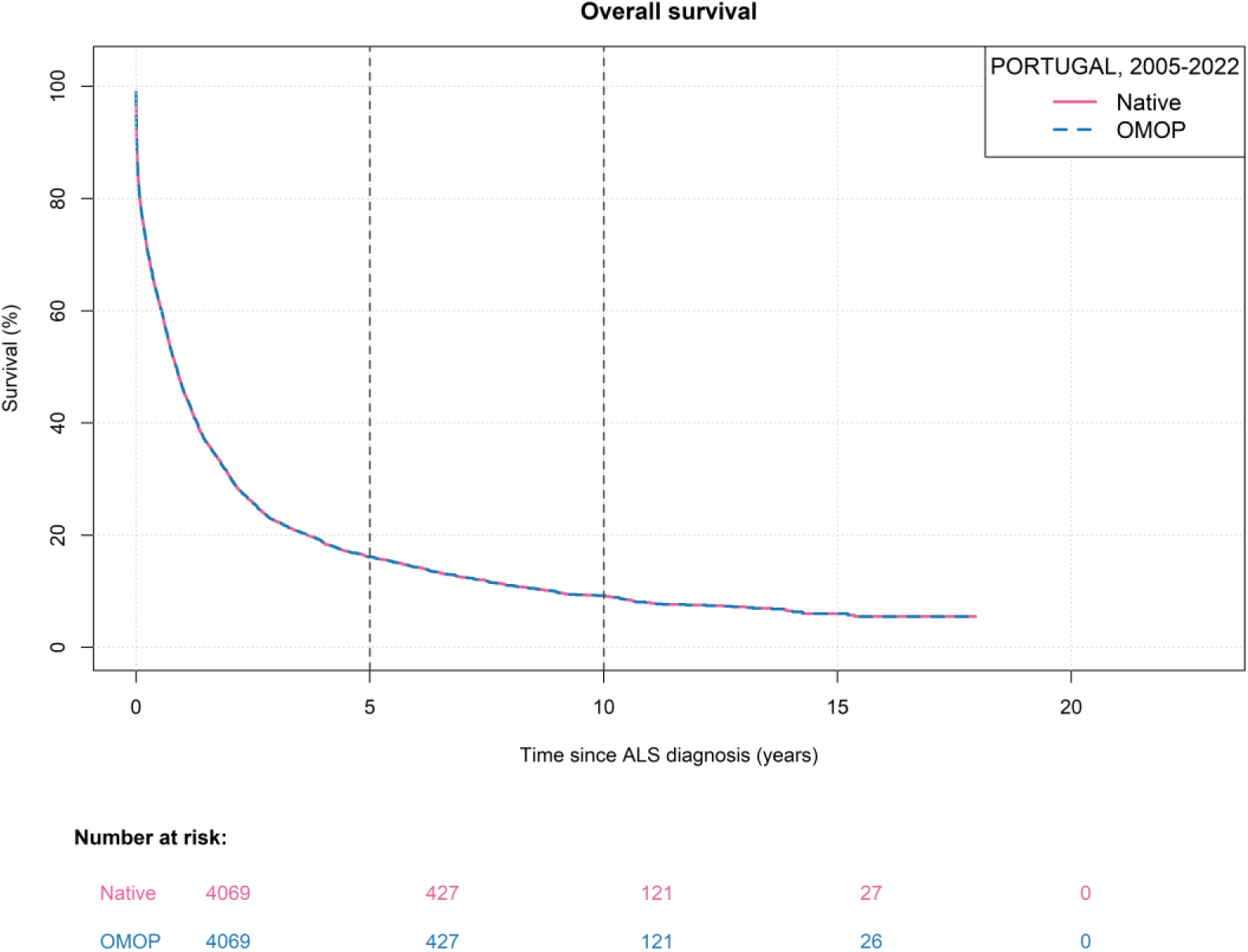
Kaplan-Meier survival curve of ALS in the Portuguese native and OMOP data.

Across all analyses, native and OMOP-based estimates showed strong agreement, supporting the validity of the harmonisation process. Observed deviations were generally small and remained within an acceptable range (5-10%).

## Discussion

In this multi-country RWD study, we assessed whether transformation of heterogeneous European RWD sources to the OMOP CDM supports semantically aligned and analytically consistent downstream analyses across countries. Using female BC and ALS as two contrasting use-case populations, we evaluated the feasibility of OMOP-based harmonisation, assessed data quality after conversion, and benchmarked OMOP-derived descriptive analyses against analyses conducted on the original native data. The study therefore addresses an important empirical question in regulatory and health-data science: whether CDM conversion improves the comparability of cross-country RWD analyses, and where residual heterogeneity persists despite harmonisation.

Overall, OMOP transformation was successfully implemented for the three participating countries and enabled the application of standardised descriptive analysis workflows across heterogeneous national data sources. The resulting OMOP databases achieved high data quality metrics, and descriptive estimates derived from OMOP-transformed data were largely concordant with estimates obtained from native-format analyses. This concordance indicates that the ETL processes preserved key epidemiological and clinical characteristics and supports the feasibility of OMOP-based harmonisation in diverse European healthcare environments. At the same time, our findings show that successful CDM conversion should not be interpreted as an automatic guarantee of cross-country comparability. Rather, there is a need for systematic quality assessment, iterative benchmarking against native data, and disease- and context-specific interpretation of harmonised data.

Heterogeneity in data completeness and granularity was observed across the countries and between the two showcased diseases, reflecting differences in healthcare organisation, coding practices, and data collection purposes. For BC, core epidemiological estimates including age, sex, incidence and mortality were broadly consistent across the participating countries and aligned with the evidence, supporting the validity of registry-based cancer data for population description [9, 10]. In particular, the stability of incidence estimates across the Nordic datasets is consistent with the well-documented burden of breast cancer in high-income settings with improved screening programmes and comprehensive cancer registration [8, 9]. However, wide variation in the availability of tumour stage and molecular characteristics mirrors known limitations in routine data capture, where clinically relevant prognostic detail is often not consistently captured in the national data sources used in this study. These findings reinforce that, while national registries reliably capture diagnoses and survival trends, more granular information required for stratified or personalised analyses — such as receptor status or detailed staging — may require linkage to additional sources of information in some settings.

The ALS analyses highlighted challenges specific to rare diseases. Variation in ALS incidence estimates across countries is broadly consistent with prior reports of higher rates in populations of European descent and likely reflect differences in diagnostic coding practices and registry coverage rather than true epidemiological differences alone [11, 12]. Despite differences in data availability and capture across countries, observed age and sex distributions were comparable across the datasets and aligned with the known age and sex distributions [31]. The inconsistent capture of ALS treatments, such as riluzole, particularly in settings where hospital-dispensed medications are not recorded at the individual level, highlights an important limitation for treatment-pattern analyses and underscores the need for disease-specific knowledge when interpreting RWD [32]. Together, these findings illustrate that while RWD are well-suited to describing the broad epidemiology and outcomes of rare diseases such as ALS, their use for more detailed clinical characterisation or treatment evaluation requires careful consideration of disease-specific pathways of care and data capture mechanisms.

Across all participating countries, linkage at the individual level within universal healthcare systems enabled longitudinal follow-up and integration of diagnoses, medication dispensing, and mortality. However, the availability of additional variables, such as socioeconomic indicators or molecular tumour markers, was often limited by country-specific data access and governance rules, which determined whether these data could be linked, accessed, or shared for the analyses. Thus, the observed data heterogeneity in RWD reflects not only differences in healthcare delivery, but also differences in the legal and institutional frameworks governing data availability and use.

A key methodological contribution of this work is the comparison of analyses conducted on native data and data transformed to the OMOP CDM. The high concordance observed between native and OMOP-based descriptive estimates is consistent with prior work demonstrating that OMOP enables population-level characterisation across heterogeneous data sources [15, 33]. However, the observed loss of breast cancer stage information in the Danish OMOP dataset highlights an important caveat: clinically relevant variables present in source registries may require additional transformation steps during the ETL process. In particular, complex variables such as cancer stage, which are often recorded through multiple registry fields or local coding conventions, may not be fully preserved through standard OMOP mappings unless explicit transformation logic is implemented [34]. Achieving concordance between native and OMOP-based results required iterative benchmarking against native analyses and refinement of ETL workflows, highlighting that harmonisation requires validation and adjustment rather than purely automated conversion [35]. For example, discrepancies observed during early comparisons of descriptive outputs were traced to differences in the interpretation of source registry variables and their corresponding OMOP mappings, which were subsequently verified and aligned to ensure consistency between native and OMOP-derived estimates. These findings underscore the importance of validation against native data and transparency around ETL decisions, particularly when OMOP-based analyses are intended to inform regulatory or public health decisions [36]. These findings are further supported by earlier European studies that have explicitly compared analyses conducted on native real-world data with those derived from OMOP-transformed datasets [37–39]. In the United Kingdom, fidelity assessments of the Clinical Practice Research Datalink (CPRD) demonstrated that conversion to OMOP preserved key population characteristics and effect estimates from established pharmaco-epidemiological studies, with discrepancies largely attributable to unmapped over-the-counter medications and local coding conventions [40]. Subsequent work using linked UK primary care, hospital, and mortality data similarly reported high concordance between cohorts defined in native formats and those derived from OMOP, with only modest deviations in selected comorbidities and medication capture [39]. Comparable observations have been reported in Germany, where multi-centre hospital implementations of OMOP showed close agreement between native and OMOP-based descriptive analyses while also documenting domain-specific information loss related to national terminologies and local documentation practices [14, 41]. Collectively, these European comparisons indicate that OMOP-based analyses can reliably reproduce population-level descriptive findings obtained from native data when ETL processes are carefully designed and validated, while also highlighting that clinically relevant detail may be attenuated without disease- and context-specific adaptations. In this regard, our observations for breast cancer staging and ALS treatment capture align closely with previously reported findings and reinforce the need for iterative benchmarking against native data when OMOP is used to support regulatory or public-health decision-making. Expert knowledge remains essential to assess fitness for purpose, particularly when using these data for comparative analyses meant to provide evidence about safety and effectiveness of treatments [11].

This study has several strengths, including its multi-country design with different health systems, inclusion of both rare disease and prevalent malignancy, and systematic comparison of native and harmonised analyses. However, also limitations must be acknowledged. Misclassification and missing data remain inherent to routine healthcare data; and access delays limited the completeness of cross-country comparisons. Importantly, these limitations reflect structural characteristics of current RWD ecosystems rather than shortcomings of individual datasets.

In conclusion, this study shows that OMOP CDM conversion can support semantically aligned, reproducible, and analytically consistent cross-country analyses of European RWD when accompanied by rigorous quality assessment and iterative validation against native data. However, OMOP conversion does not by itself eliminate heterogeneity in source-data provenance, clinical practice, coding systems, completeness, or governance constraints. Cross-country comparability remains a study-specific empirical property that must be demonstrated rather than assumed. Robust use of harmonised RWD for regulatory, HTA, or public-health evidence generation therefore requires transparent ETL documentation, native-data benchmarking, and close collaboration between clinical, epidemiological, and data-science experts.

## Data Availability

The data used in this study originate from the registries and databases in Finland, Denmark, and Portugal and are subject to all local legal and data governance restrictions. As a result, the datasets are not publicly available and cannot be shared by the authors. Access to these data may be granted by the respective national data custodians following appropriate approvals in accordance with national regulations.

## Code Availability

The code used for the data transformation and analyses can be accessed via https://github.com/mohaborageh/OMOP-Analysis. The code is provided to document the analytical workflow used in the article and to support transparency and reproducibility where possible, within the constraints of the applicable data policies.

## Funding

The research leading to these results has received funding from the European Community’s Horizon Europe Programme under grant agreement no. 101095353 (Real4Reg). Views and opinions expressed are those of the authors only and do not necessarily reflect those of the European Union or the European Commission. Neither the European Union nor the granting authority can be held responsible for them. The funding organisations had no role in the design and conduct of the study; collection, management, analysis and interpretation of the data; preparation, review or approval of the manuscript; and decision to submit the manuscript for publication.

## Author Contributions

Acquisition of funding: AMT, SH, VE, HF, BH; conceptualisation: VE, HF; data analysis: MA, MRKH, IB, EHP, ML, BR, TV, CS, CV; drafting of manuscript: MA, MRKH; review and editing of manuscript: CF, CS, EHP, JF, CB, BR, AP, TV, SH, AMT, HF, VE, BH.

## Acknowledgements

We thank EUpALS (Dirk De Valck and Evy Reviers) and the rest of the Real4Reg consortium for helpful discussions and intellectual input to this paper.

## Declarations

The authors declare no conflict of interest.

## Clinical Trial Number

Not applicable.

## Notes

### Competing Interest Statement

The authors have declared no competing interest.

### Author Declarations

In Denmark and Finland the ethics approval or informed consent were not required as the study was conducted under the legislation on secondary use of health and social data. Research team had access to pseudonymised data and study participants were not contacted All analyses were conducted within secure national data environments (Danish Health Data Authority, National Genome Center in Denmark and Findata in Finland. The Portuguese Ethics Committee for Clinical Research (CEIC) approved the study plan (Deliberation no. 2024-RP-04-13, 12 April 2024). All data were pseudonymised before access by the research team, and all analyses were conducted within secure national data environments at INFARMED, I.P. Access, in all countries was limited to authorised researchers in accordance with national regulations. Only aggregated results were reported, following national disclosure rules to prevent re-identification of individuals and privacy breaches. Minimum cell-count thresholds were applied (Finland: <;3; Denmark: <5; Portugal: <10;), with Denmark additionally rounding person counts to the nearest five in the native analysis results. All derived descriptive statistics, incidence and prevalence measures, and mortality estimates were calculated from unrounded source data.

## References

1. Prilla S, Groeneveld S, Pacurariu A, Restrepo-Méndez MC, Verpillat P, Torre C, et al. Real-World Evidence to Support EU Regulatory Decision Making—Results From a Pilot of Regulatory Use Cases. Clin Pharma and Therapeutics. 2024;116:1188–97. 10.1002/cpt.3355.

2. European Medicines Agency. Reflection paper on establishing efficacy based on single-arm trials submitted as pivotal evidence in a marketing authorisation application. Amsterdam: European Medicines Agency; 2024.

3. European Medicines Agency. Draft concept paper on the development of a reflection paper on the use of external controls for evidence generation for regulatory decision-making. Amsterdam: European Medicines Agency; 2025.

4. Mack C, Christian J, Brinkley E, Warren EJ, Hall M, Dreyer N. When Context Is Hard to Come By: External Comparators and How to Use Them. Drug Inf J. 2019;:216847901987867. 10.1177/2168479019878672.

5. Khozin S, Dreyer NA, Galante D, Liu R, Neumann P, Nussbaum N, et al. Real-World Evidence Acceptability and Use in Breast Cancer Treatment Decision-Making in the United States: Call-to-Action from a Multidisciplinary Think Tank. Adv Ther. 2025;42:2973–87. 10.1007/s12325-025-03201-y.

6. Makady A, De Boer A, Hillege H, Klungel O, Goettsch W. What Is Real-World Data? A Review of Definitions Based on Literature and Stakeholder Interviews. Value in Health. 2017;20:858–65. 10.1016/j.jval.2017.03.008.

7. Makady A, Van Veelen A, Jonsson P, Moseley O, D’Andon A, De Boer A, et al. Using Real-World Data in Health Technology Assessment (HTA) Practice: A Comparative Study of Five HTA Agencies. PharmacoEconomics. 2018;36:359–68. 10.1007/s40273-017-0596-z.

8. European Commission. Development, optimisation and implementation of artificial intelligence methods for real-world data analyses in regulatory decision-making and health technology assessment along the product lifecycle. Brussels: European Commission; 2024. 10.3030/101095353.

9. Bray F, Laversanne M, Sung H, Ferlay J, Siegel RL, Soerjomataram I, et al. Global cancer statistics 2022: GLOBOCAN estimates of incidence and mortality worldwide for 36 cancers in 185 countries. CA A Cancer J Clinicians. 2024;74:229–63. 10.3322/caac.21834.

10. Kim J, Harper A, McCormack V, Sung H, Houssami N, Morgan E, et al. Global patterns and trends in breast cancer incidence and mortality across 185 countries. Nat Med. 2025;31:1154–62. 10.1038/s41591-025-03502-3.

11. Xu L, Liu T, Liu L, Yao X, Chen L, Fan D, et al. Global variation in prevalence and incidence of amyotrophic lateral sclerosis: a systematic review and meta-analysis. J Neurol. 2020;267:944–53. 10.1007/s00415-019-09652-y.

12. Wolfson C, Gauvin DE, Ishola F, Oskoui M. Global Prevalence and Incidence of Amyotrophic Lateral Sclerosis: A Systematic Review. Neurology. 2023;101. 10.1212/WNL.0000000000207474.

13. Voss EA, Blacketer C, Van Sandijk S, Moinat M, Kallfelz M, Van Speybroeck M, et al. European Health Data & Evidence Network—learnings from building out a standardized international health data network. Journal of the American Medical Informatics Association. 2023;31:209–19. 10.1093/jamia/ocad214.

14. Maier C, Lang L, Storf H, Vormstein P, Bieber R, Bernarding J, et al. Towards Implementation of OMOP in a German University Hospital Consortium. Appl Clin Inform. 2018;09:054–61. 10.1055/s-0037-1617452.

15. Hripcsak G, Duke JD, Shah NH, Reich CG, Huser V, Schuemie MJ, et al. Observational Health Data Sciences and Informatics (OHDSI): Opportunities for Observational Researchers. Stud Health Technol Inform. 2015;216:574–8.

16. Ganna A, Carracedo A, Christiansen CF, Di Angelantonio E, Dykstra PA, Dzhambov AM, et al. The European Health Data Space can be a boost for research beyond borders. Nat Med. 2024;30:3053–6. 10.1038/s41591-024-03246-6.

17. Arlett P, Kjær J, Broich K, Cooke E. Real-World Evidence in EU Medicines Regulation: Enabling Use and Establishing Value. Clin Pharma and Therapeutics. 2022;111:21–3. 10.1002/cpt.2479.

18. Voss EA, Shoaibi A, Yin Hui Lai L, Blacketer C, Alshammari T, Makadia R, et al. Contextualising adverse events of special interest to characterise the baseline incidence rates in 24 million patients with COVID-19 across 26 databases: a multinational retrospective cohort study. eClinicalMedicine. 2023;58:101932. 10.1016/j.eclinm.2023.101932.

19. Schmidt M, Schmidt SAJ, Adelborg K, Sundbøll J, Laugesen K, Ehrenstein V, et al. The Danish health care system and epidemiological research: from health care contacts to database records. CLEP. 2019;Volume 11:563–91. 10.2147/CLEP.S179083.

20. de Almeida Simoes J, Augusto GF, Fronteira I, Hernandez-Quevedo C. Portugal: Health System Review. Health Syst Transit. 2017;19:1–184.

21. Keskimaki I, Tynkkynen L-K, Reissell E, Koivusalo M, Syrja V, Vuorenkoski L, et al. Finland: Health System Review. Health Syst Transit. 2019;21:1–166.

22. Observational Health Data Sciences and Informatics (OHDSI). OMOP Common Data Model v5.4. Available at: https://ohdsi.github.io/CommonDataModel/cdm54.html. Accessed May 26, 2026.

23. Schmidt M, Schmidt SAJ, Sandegaard JL, Ehrenstein V, Pedersen L, Sørensen HT. The Danish National Patient Registry: a review of content, data quality, and research potential. CLEP. 2015;:449. 10.2147/CLEP.S91125.

24. Kurki MI, Karjalainen J, Palta P, Sipilä TP, Kristiansson K, Donner KM, et al. FinnGen provides genetic insights from a well-phenotyped isolated population. Nature. 2023;613:508–18. 10.1038/s41586-022-05473-8.

25. Gracia-Tabuenca J, Koskenvesa P, Tajanen P, Kukkurainen S, Klingstedt G. FinOMOP – a population-based data network. OHDSI; 2023.

26. European Commission. Statistical Office of the European Union. Revision of the European Standard Population: report of Eurostat’s task force : 2013 edition. LU: Publications Office; 2013. 10.2785/11470.

27. R Core Team. R: A language and environment for statistical computing. R Foundation for Statistical Computing; 2025.

28. Facile R, Muhlbradt EE, Gong M, Li Q, Popat V, Pétavy F, et al. Use of Clinical Data Interchange Standards Consortium (CDISC) Standards for Real-world Data: Expert Perspectives From a Qualitative Delphi Survey. JMIR Med Inform. 2022;10:e30363. 10.2196/30363.

29. Gjerstorff ML. The Danish Cancer Registry. Scand J Public Health. 2011;39 7_suppl:42–5. 10.1177/1403494810393562.

30. Leinonen MK, Miettinen J, Heikkinen S, Pitkäniemi J, Malila N. Quality measures of the population-based Finnish Cancer Registry indicate sound data quality for solid malignant tumours. European Journal of Cancer. 2017;77:31–9. 10.1016/j.ejca.2017.02.017.

31. Riva N, Domi T, Pozzi L, Lunetta C, Schito P, Spinelli EG, et al. Update on recent advances in amyotrophic lateral sclerosis. J Neurol. 2024;271:4693–723. 10.1007/s00415-024-12435-9.

32. Feldman EL, Goutman SA, Petri S, Mazzini L, Savelieff MG, Shaw PJ, et al. Amyotrophic lateral sclerosis. The Lancet. 2022;400:1363–80. 10.1016/S0140-6736(22)01272-7.

33. Voss EA, Makadia R, Matcho A, Ma Q, Knoll C, Schuemie M, et al. Feasibility and utility of applications of the common data model to multiple, disparate observational health databases. Journal of the American Medical Informatics Association. 2015;22:553–64. 10.1093/jamia/ocu023.

34. Blacketer C, Defalco FJ, Ryan PB, Rijnbeek PR. Increasing trust in real-world evidence through evaluation of observational data quality. Journal of the American Medical Informatics Association. 2021;28:2251–7. 10.1093/jamia/ocab132.

35. Kahn MG, Callahan TJ, Barnard J, Bauck AE, Brown J, Davidson BN, et al. A Harmonized Data Quality Assessment Terminology and Framework for the Secondary Use of Electronic Health Record Data. eGEMs. 2016;4:18. 10.13063/2327-9214.1244.

36. Zoch M, Gierschner C, Peng Y, Gruhl M, Leutner LizA, Sedlmayr M, et al. Adaption of the OMOP CDM for Rare Diseases. In: Mantas J, Stoicu-Tivadar L, Chronaki C, Hasman A, Weber P, Gallos P, et al., editors. Studies in Health Technology and Informatics. IOS Press; 2021. 10.3233/SHTI210136.

37. Hunt NB, Souverein P, Bazelier M, Barclay N, Delmestri A, Sturkenboom M, et al. Implementation of OMOP and ConcePTION Common Data Models in CPRD GOLD: Risk of Bleeding and Cardiovascular Outcomes From Anticoagulant Use. Clin Pharma and Therapeutics. 2026;:cpt.70242. 10.1002/cpt.70242.

38. Biedermann P, Ong R, Davydov A, Orlova A, Solovyev P, Sun H, et al. Standardizing registry data to the OMOP Common Data Model: experience from three pulmonary hypertension databases. BMC Med Res Methodol. 2021;21:238. 10.1186/s12874-021-01434-3.

39. Papez V, Moinat M, Payralbe S, Asselbergs FW, Lumbers RT, Hemingway H, et al. Transforming and evaluating electronic health record disease phenotyping algorithms using the OMOP common data model: a case study in heart failure. JAMIA Open. 2021;4:ooab001. 10.1093/jamiaopen/ooab001.

40. Matcho A, Ryan P, Fife D, Reich C. Fidelity Assessment of a Clinical Practice Research Datalink Conversion to the OMOP Common Data Model. Drug Saf. 2014;37:945–59. 10.1007/s40264-014-0214-3.

41. Hechtel N, Apfel-Starke J, Köhler S, Fradziak M, Schönfeld N, Steinmeyer J, et al. Harmonisation of German Health Care Data Using the OMOP Common Data Model – A Practice Report. In: Mantas J, Gallos P, Zoulias E, Hasman A, Househ MS, Charalampidou M, et al., editors. Studies in Health Technology and Informatics. IOS Press; 2023. 10.3233/SHTI230485.

