## Supplementary Materials for "Does OMOP CDM Conversion Improve Cross-Country Comparability of Real-World Data? A Benchmark Study in Breast Cancer and Amyotrophic Lateral Sclerosis"

### Harmonising European Real-World Data for Regulatory Evidence Generation: OMOP-Based Analyses of Breast Cancer and Amyotrophic Lateral Sclerosis

### Supplementary Materials

Mohamed Aborageh[1,2,†*],Monika Roberta Korcinska Handest[3,†], Istvan Bakos[4],Blair Rajamaki[5], Célia Silva[6], Erzsébet Horváth-Puhó[4], Liisa Pylkkänen[7], Catarina Venda[6], Manuel Lentzen[1,2], Cornelia Becker[8], Joana Fernandes[6], Anne Paakinaho[5], Thuan Vo[5], Britta Haenisch[8,11,12], Sirpa Hartikainen[5], Anna-Maija Tolppanen[5], Claudia Furtado[6,9], Holger Fröhlich[1,2,10,†*], Vera Ehrenstein[4,†*]

Affiliations:

[1] Fraunhofer Institute for Algorithms and Scientific Computing SCAI, Department of Biomedical AI & Data Science, Sankt Augustin, Germany

[2] University of Bonn, Bonn-Aachen International Center for IT (b-it), Bonn, Germany

[3] Danish Medicines Agency, Data and Data Analytic Center (DAD), Copenhagen, Denmark

[4] Aarhus University and Aarhus University Hospital, Department of Clinical Epidemiology, Center for Population Medicine, Aarhus, Denmark

[5] University of Eastern Finland, Faculty of Health Sciences, School of Pharmacy, Kuopio, Finland

[6] INFARMED – National Authority of Medicines and Health Products, I.P., Lisbon, Portugal

[7] University of Turku, Department of Oncology, Turku, Finland

[8] Federal Institute for Drugs and Medical Devices, Research Division, Bonn, Germany

[9] NOVA University Lisbon, NOVA National School of Public Health, Lisbon, Portugal

[10] University of Bonn, University Hospital Bonn, Institute for Digital Medicine, Bonn, Germany

[11] German Center for Neurodegenerative Diseases (DZNE), Bonn, Germany

[12] University of Bonn, University Hospital Bonn, Center for Translational Medicine, Bonn, Germany

[†] These authors contributed equally to this work

#### Supplementary Material 1: Selected Common Data Model (CDM) Tables and Descriptions

**Person:** demographics of each person (e.g., birth year, gender, race)

**Death:** death information including date and cause

**Visit_occurrence:** records of each healthcare visit or encounter

**Condition_occurrence:** diagnoses or medical conditions assigned to a person

**Drug_exposure:** medications prescribed, dispensed or administered

**Procedure_occurrence:** surgical or medical procedures performed. Would also include cancer specific procedures including chemotherapy regimens

**Measurement:** laboratory tests or other measurements with results

**Observation_period:** time span in which data are available for a person

**Concept:** core vocabulary table where each unique concept has a concept ID

**Concept_relationship:** links between concepts (e.g., mappings)

**Vocabulary:** metadata about each vocabulary (e.g., SNOMED, LOINC, ICD-10)

#### Supplementary Material 2: Used Vocabulary and their Mapping

**Denmark:** The Danish Healthcare Classification System (Danish Sundhedsvæsenets Klassifikationssystem SKS) is used to standardise clinical coding across Danish healthcare systems and is structured as follows:

- B: Danish classification of non-surgical treatment, care and prophylaxis given in hospitals
- D: Danish version of the ICD-10. Contains codes for diagnoses, symptoms and health problems as recorded in hospitals
- K: Danish version of the “NOMESCO Classification of Surgical Procedures” (NSCP) and contains codes for surgical procedures
- ATC system: used for the classification of drugs in several Danish Registries

ICD-10 and ATC codes were mapped using pre-existing concept maps obtained from Athena. According to the suggestion of a Real4Reg Danish partner, Aarhus University

(AU) we used the mappings developed by FinOMOP to map to NSCP. This decision was taken due to the overlap of NSCP codes in Finland and Denmark. For non-surgical treatment, care and prophylaxis (codes with leading B), a manual mapping was necessary as they included country specific concepts that are not yet mapped to standardised codes.

**Finland:** For Finland, we used vocabulary files generated as part of the FinOMOP and FinnGen projects (https://www.ohdsi-europe.org/index.php/national-nodes/finland). These vocabulary files build on top of the Athena vocabularies and include e.g. certain ICD codes that are not part of the ICD-10 or ICD-10-CM version.

- ICD-9 and ICD-10 (with extension): The ICD classification system is used for diagnoses, symptoms, and health problems. Here, different versions are used, including the extensions provided by FinOMOP.
- ICD-O-3: WHO-endorsed coding system designed for neoplasms, capturing topography and morphology
- NSCP: Nordic classification of surgical procedures
- ATC system: used for the classification of drugs

Nordic Product Numbers (VNR): Six-digit identification codes that are assigned to drugs with marketing authorization to allow reliable identification and verification of a specific drug package.

Mapping of the native codes was done using the provided vocabulary files. For certain cases (e.g. socioeconomic codes or missing VNRs), a custom vocabulary was defined with new concept IDs. For the VNR extension, we used the associated ATC codes, to map the native VNR to a RxNorm concept if that information was available for the given ATC code.

**Portugal:** Most Portuguese registries use vocabularies that are included in Athena; hence the use of external mapping projects was not necessary for most coding systems.

- ICD9-CM and ICD10-CM: The ICD classification system is used for diagnoses, symptoms and health problems for inpatient and outpatient care in hospitals.
- ICD10: The ICD classification system used for death causes.
- ICD-9-Vol3 and ICD-10-PCS: International Classification of Diseases, Procedure Coding System, used to code procedures in the hospital settings.
- ICD-9-Proc and ICD-10-PCS: The ICD classification system for procedure coding. Used to code inpatient procedures only.
- ATC system: used for classification of drugs
- Medicine Product Number (NRM: "Número de Registo de Medicamento"): Seven-digit identification codes that are assigned to medicines with marketing authorization to allow reliable identification and verification of a specific package. The NRM is a unique and specific code for each medicine and pharmaceutical presentation.

Mapping of the native ICD and ATC codes was done using the provided vocabulary files. With the NRM code it was possible to map to the associated ATC codes. In addition to the systems mentioned above, the International Classification of Primary Care (ICPC-2) classification system for diagnoses, symptoms, health problems and procedures in primary care settings is used. Since this is not represented in Athena, custom concepts will be added to represent the data in the OMOP databases.

#### Supplementary Material 3: Analytical Tools and Data Quality Assessment

Data extraction, transformation and analysis were conducted within the OMOP Common Data Model Framework using structured query language (SQL) and R (R Foundation for Statistical Computing, Vienna, Austria). Database interactions were performed using *DBI*, *RPostgres*, and *dbplyr*, enabling efficient querying and manipulation of OMOP tables in PostgreSQL and DuckDB environments. Data processing and cohort construction were implemented using the *tidyverse* suite (including *dbplyr*) and *lubridate* for temporal handling. Descriptive analyses and visualisations were conducted using *ggplot2*.

Data quality was systematically assessed using the OHDSI Data Quality Dashboard, which applies standardised conformance, completeness, and plausibility checks across the dataset. Additional validation procedures included consistency checks against source data, iterative benchmarking of descriptive estimates between native and OMOP-converted datasets, and targeted inspection of key variables to ensure fidelity of the transformation process.

**Supplementary** Table 1. Overview of the country-specific real world data sources used in the Real4Reg project

| **Country, population size, coverage (%)** | **Data source: local name** | **Data source: English name** | **Data provenance** | **Source population** | **Prompt: event causing a record in the data source** | **Content in Real4Reg** | **Data dictionaries**<br>**Data types** |
|---|---|---|---|---|---|---|---|
| DENMARK 5.7 mio 100% coverage | CPR-registeret | CPR Registry | Total population registry | All residents | Birth | Personal identifier (pseudonymised) | Local structured codes |
| | | | | | Immigration | Sex | Dates |
| | | | | | | Age | |
| | | | | | | Residence | |
| | | | | | | Vital status | |
| | | | | | | Migrations | |
| | Lægemiddelstatistikregisteret | National Prescription Registry | Community pharmacies | All residents | Pharmacy dispensing of a prescribed medicine | Active substance | ATC |
| | | | | | | Date of dispensing | Dates |
| | | | | | | Amount dispensed | Alphanumeric data |
| | Landspatientregisteret | National Patient Registry | Private and public ambulatory and inpatient hospital departments | All residents | Hospitalisation | Discharge diagnoses | ICD-10 |
| | | | | | Visit to a hospital specialist clinic | Surgeries | NOMESCO |
| | | | | | | Hospital treatments | Local structured codes |
| | | | | | | Diagnostic procedures | |
| | Patologiregisteret | Pathology Registry | Pathology departments | All residents | Pathology report | Molecular/genetic profile (breast cancer) | SNOMED |
| | Cancerregisteret | Cancer Registry | Oncology departments | All residents | Diagnosis of new primary malignancy | Diagnosis | ICD-10 |
| | | | | | | Morphology | ICD-O |
| | | | | | | Stage at diagnosis | TNM |
| | Dødsårsagsregisteret | Cause of Death Registry | Death certificates | All residents | Death certificate | Underlying cause of death | ICD-10 |
| FINLAND 5.6 mio 100% coverage | Digi- ja väestötietoviraston varmennepalveluista (DVV) väestötietojärjestelmä | Population information system | Total population registry | All residents | Birth | Personal identifier (pseudonymised) | Local structured codes |
| | | | | | Immigration, Emigration | Sex | Dates |
| | | | | | | Age | |
| | | | | | | Residence | |
| | | | | | | Vital status | |

| | | | | | | | |
|---|---|---|---|---|---|---|---|
| | | | | | | Migrations | |
| | Lääketoimitukset Kanta-Reseptikeskus | Kanta electronic prescriptions dispensings | Community pharmacies | All residents | Pharmacy dispensing of a prescribed medicine | Active substance Drug name | ATC |
| | | | | | | Date of dispensing | Dates |
| | | | | | | Amount dispensed | Alphanumeric data |
| | | | | | | Strength | |
| | Sairausvakuutuksesta korvattavat lääketoimitukset | Dispensed medicines reimbursable under the National Health Insurance scheme | Community pharmacies | All residents | Pharmacy dispensing of a prescribed medicine reimbursable under the National Health Insurance scheme | Active substance Drug name | ATC |
| | | | | | | Date of dispensing | Dates |
| | | | | | | Amount dispensed | Alphanumeric data |
| | | | | | | Strength | |
| | Lääkekorvausoikeudet | Entitlements to reimbursement of pharmaceutical expenses | Private and public primary and secondary healthcare | All residents | Reimbursement of pharmaceutical expenses | Comorbidity information, Reimbursement code | Dates |
| | | | | | | | Local structured codes |
| | | | | | | | ICD-9, ICD-10 |
| | Terveydenhuollon hoitoilmoitusrekisteri | Care Register for Health Care | Private and public inpatient | All residents | Hospitalization | Diagnosis | Dates |
| | | | | | | Procedures | ICD-10, ICD-9 |
| | | | | | | Required level of assistance at discharge | NOMESCO |
| | Perusterveydenhuollon avohoidon hoitoilmoitusrekisteri AvoHILMO | Register of Primary Health Care Visits | Public healthcare | All residents | Primary Health Care Visits | General healthcare outpatient visits Diagnosis Procedure | Dates |
| | | | | | | | ICD-10 |
| | | | | | | | NOMESCO |
| | | | | | | | ICPC-2 |
| | Kuolemansyiden tutkimusaineisto | Causes of Death Register | Death certificates | All residents | Death certificates | Death dates, causes, how the cause was ascertained | Dates |
| | | | | | | | Local structured codes |
| | | | | | | | ICD-10 |
| | Syöpärekisteri | Cancer Registry | Oncology clinics | All residents | Cancer diagnosis | Cancer diagnosis | ICD-10 |
| | | | | | | | ICD-O-3 |
| | | | | | | | Dates |
| PORTUGAL 10.3 mio 100% coverage | Registo Nacional de Utentes | National User Register | National Health Service (NHS) | All residents | Issuance of the Citizen Card (automatically), or manually by | Personal identifier (pseudonymised) | Local structured codes |
| | | | | | | Date of birth | Dates |

| | | | | | | | |
|---|---|---|---|---|---|---|---|
| | | | | | healthcare administrative staff (in some cases) | Date of death | |
| | | | | | | Sex | |
| | Base de Dados Nacional de Prescrição (BDNP) | National Prescription Database | National Health System (NHS) | All residents | Pharmacy dispensing of a prescribed medicine reimbursed by the NHS | Medicine Product Number | Local structured codes |
| | | | | | | Date of dispensing | Dates |
| | | | | | | Amount dispensed | ATC |
| | | | | | | | Alphanumeric data |
| | Base de Dados de Morbilidade Hospitalar (BDMH) | Hospital Morbidity Database | Public hospital care | All residents | Hospital visit | Diagnosis | Local structured codes |
| | | | | | | Procedures | Dates |
| | | | | | | | ICD-9-CM |
| | | | | | | | ICD-10-CM/PCS |
| | SIM@SNS — Sistema de Informação e Monitorização do SNS | NHS Information and Monitoring System | NHS -Primary Care | All residents | Primary health care visit | Diagnosis | Local structured codes |
| | | | | | | Problems | Dates |
| | | | | | | Procedures | ICPC-2 |
| | Registo Oncológico Nacional (RON) | Cancer register database | Oncology services of the hospitals | All residents | Cancer diagnosis | Diagnosis | Local structured codes |
| | | | | | | Morphology | Dates |
| | | | | | | Topography | ICD-O-3 |
| | | | | | | Treatments | |
| | | | | | | Procedures | |
| | Sistema de Informação dos Certificados de Óbito (SICO) | Death register database | Death certificates | All residents | Death certificates | Death date | Dates |
| | | | | | | Death cause | ICD-10 |

**Supplementary Table 2: Demographic and clinical characteristics of women with incident breast cancer in participating countries (Denmark and Finland, 2000–2021; Portugal, 2005–2022)**

| Characteristics | Denmark | | Finland | | Portugal | |
|---|---|---|---|---|---|---|
| | NATIVE | OMOP | NATIVE | OMOP | NATIVE | OMOP |
| **Women** | 97550 (100%) | 97440 (100%) | 87782 (100%) | 87782 (100%) | 98345 (100%) | 97546(100%) |
| **Age, years** | | | | | | |
| 18 - 34 | 1525 (2%) | 1527 (2%) | 994 (1%) | 994 (1%) | 2465 (3%) | 2445 (3%) |
| 35 - 44 | 7240 (7%) | 7241 (7%) | 5191 (6%) | 5191 (6%) | 12065( 12%) | 11986 (12%) |
| 45 - 54 | 18745 (19%) | 18746 (19%) | 18198 (21%) | 18198 (21%) | 22905(23%) | 22749 (23%) |
| 55 - 64 | 25330 (26%) | 25331 (26%) | 25137 (29%) | 25137 (29%) | 22803 (23%) | 22583 (23%) |
| 65 - 74 | 23490 (24%) | 23488 (24%) | 20319 (23%) | 20319 (23%) | 20306 (21%) | 20135 (21%) |
| >=75 | 21220 (22%) | 21220 (22%) | 17943 (20%) | 17943 (20%) | 17801(18%) | 17648 (18%) |
| Median (quartiles) | 64 (53-73) | 63 (53-73) | 62 (53-72) | 62 (53-72) | 60( 49-71) | 60(49-71) |
| **Year of diagnosis** | | | | | | |
| 2000 - 2010 (2005-2013 PT) | 46705 (48%) | 46705 (48%) | 40993 (47%) | 40993 (47%) | 44214 (45%) | 44092 (45%) |
| **Pregnancy** | 145 (<1%) | 144 (<1%) | 80 (<1%) | 80 (<1%) | 137 (<1%) | 396((<1%) |
| **Parity**[1*] | | | | | | |
| 0 | 60 (<1%) | 136 (<1%) | NA | NA | NA | NA |
| 1 | 60 (<1%) | 88 ((<1%) | 1346 (2%) | 1346 (2%) | 487(<1%) | 580(1%) |
| 2 | 25 (<1%) | 31 (<1%) | NA | NA | 21((<1%) | 27 (<1%) |
| NA | 97410 (100%) | 97298 (100%) | NA | NA | NA | NA |
| **Comorbidities** [1] | | | | | | |
| Psychiatric disorders [2] | 740 (1%) | 845 (1%) | 3255 (4%) | 3255 (4%) | 337(<1%) | 314(<1%) |

| | | | | | | |
|---|---|---|---|---|---|---|
| Dementia | 1210 (1%) | 1431 (2%) | 2619 (3%) | 2742 (3%) | 655(1%) | 821(1%) |
| Cardiovascular diseases[3] | 7840 (8%) | 7507 (8%) | 22741 (26%) | 22742 (26%) | 8131(8%) | 8995(9%) |
| **Medication use** [4] | | | | | | |
| Systemic contraceptives[1] | 7190 (7%) | 7188 (7%) | 2464 (3%) | 2464 (3%) | 3261(8%) | 3210(8%) |
| Systemic contraceptives: After Index Date | 865 (1%) | 888 (1%) | 337 (<1%) | 337 (<1%) | 289(1%) | 298(1%) |
| HRT usage | 16455 (17%) | 16137 (17%) | 24739 (28%) | 24740 (28%) | 4535(2%) | 4445(11%) |
| HRT usage: After Index Date | 3845 (4%) | 4744 (5%) | 2295 (3%) | 2296 (3%) | 775(0%) | 801(2%) |
| **Receptor status** | | | | | | |
| Androgen receptor positive | 65 ((<1%) | 71 (<1%) | NA | NA | NA | NA |
| Androgen receptor negative | 10 (<1%) | 12 (<1%) | NA | NA | NA | NA |
| Estrogen receptor positive | 16020 (16%) | 17310 (18%) | NA | NA | 32997 (34%) | 33327 (34%) |
| Estrogen receptor negative | 4330 (4%) | 4697 (5%) | NA | NA | 6556 (7%) | 6577 (7%) |
| Progesterone receptor positive | 3075 (3%) | 3312 (3%) | NA | NA | 61145 (62%) | 62123 (64%) |
| Progesterone receptor negative | 2095 (2%) | 2259 (2%) | NA | NA | 10397 (11%) | 10434 (11%) |
| HER2 receptor normal expression | 16615 (17%) | 17769 (18%) | NA | NA | 6840 (7%) | 6858 (7%) |
| HER2 receptor borderline expression | 2310 (2%) | 2467 (3%) | NA | NA | NA | NA |
| HER2 receptor overexpression | 2550 (3%) | 2791 (3%) | NA | NA | NA | NA |
| HER2 negative | 30 (<1%) | 0 (0%) | NA | NA | 29008 (29%) | 29295 (30%) |
| HER2 expression 1+ (>10%) | 15 (<1%) | 0 (0%) | NA | NA | NA | NA |
| HER2 expression 2+ | 0 (0%) | 0 (0%) | NA | NA | NA | NA |
| HER2 expression 3+ | 5 (<1%) | 0 (0%) | NA | NA | NA | NA |
| BRCA1 gene status normal | 50 (<1%) | 0 (0%) | NA | NA | NA | NA |
| BRCA1 gene status other | 30 (<1%) | 0 (0%) | NA | NA | NA | NA |
| BRCA2 gene status normal | 55 (<1%) | 0 (0%) | NA | NA | NA | NA |
| BRCA2 gene status other** | 20 (<1%) | 0 (0%) | NA | NA | NA | NA |
| **Signals for disease treatment**[5] | | | | | | |

| | | | | | | |
|---|---|---|---|---|---|---|
| Radiotherapy | 56755 (58%) | 47898 (49%) | 21490 (24%) | 21490 (24%) | 28816 (29%) | 28816 (30%) |
| Chemotherapy | 36370 (37%) | 31228 (32%) | 21735 (25%) | 21735 (25%) | 57193 (58%) | 57020 (58%) |
| Total or partial lymphadenectomy | 57665 (59%) | 50929 (52%)) | 33442 (38%) | 33442 (38%) | 37194 (38%) | 35222 (36%) |
| Mastectomy | 33155 (34%) | 29851 (31%) | 36994 (42%) | 36994 (42%) | 67769 (69%) | 63058 (65%) |
| Breast conservation surgery | 47265 (48%) | 41679 (43%) | 42124 (48%) | 42124 (48%) | 8658 (9%) | 8238 (8%) |
| Hormonal therapy for breast cancer | 36420 (37%) | 33638 (34%) | 58188 (66%) | 58188 (66%) | 30454(31%) | 31615 (32%) |
| HER directed treatments [6] | 9400 (10%) | 7637 (8%) | 4750 (5%) | 4764 (5%) | 6244(6%) | 6225 (6%) |
| **Signals for disease progression** [7] | | | | | | |
| **Stage at diagnosis** [8]*** | | | | | | |
| TNM Stage: 1 | 42845 (44%) | 37968 (44%) | NA | NA | 33748(34%) | 32600 (33%) |
| TNM Stage: 2 | 23320 (24%) | 21422 (25%) | NA | NA | 29650 (30%) | 29407 (30%) |
| TNM Stage: 3 | 2875 (3%) | 4161 (5%) | NA | NA | 12088 (12%) | 12004 (12%) |
| TNM Stage: NA | 28515 (29%) | 22131 (26%) | NA | NA | 16925 (17%) | 15348 (16%) |
| Finland Stage: 1 localised | NA | NA | 40241 (46%) | 40240 (46%) | NA | NA |
| Finland Stage: 2 Non-localized | NA | NA | 29593 (34%) | 29600 (34%) | NA | NA |
| Finland Stage: NA | NA | NA | 17948 (20%) | 17942 (20%) | NA | NA |
| **Primary tumour size** *** | | | | | | |
| No growth beyond primary cells | 5795 (6%) | 7841 (6%) | 100 (0%) | 100 (<1%) | 567 (1%) | 561(1%) |
| Tumour growth beyond primary cells | 76330 (78%) | 72638 (78%) | 20160 (98%) | 20163 (98%) | 81019 (83%) | 78466 (80%) |
| Undefined growth | 15425 (16%) | 5203 (16%) | 39 (<1%) | 41 (<1%) | 15585 (16%) | 15582 (16%) |
| **Metastases at diagnosis** *** | | | | | | |
| Metastasis - spread to lymph nodes | 30155 (31%) | 28627 (30%) | 8551 (42%) | 8552 (43%) | 32631 (33%) | 31736 (33%) |
| Metastasis - spread to other organs | 3475 (4%) | 5341 (6%) | 300 (1%) | 300 (1%) | 5260 (5%) | 5158 (5%) |

| Other signals of progression | | | | | | |
|---|---|---|---|---|---|---|
| New primary malignancy | 8050 (8%) | 8052 (8%) | 5989 (7%) | 5989 (7%) | 11722 (12%) | 11129 (11%) |
| Breast cancer specific mortality | 16535 (17%) | 17358 (17%) | 12001 (14%) | 12001 (14%) | 8698 (16%) | 8699 (16%) |
| All-cause mortality | 32890 (34%) | 35043 (35%) | 25883 (29%) | 25874 (30%) | 24789(25%) | 24746(25%) |

Counts are masked according to each country's privacy-preserving rules
Abbreviations: NA not available
*Number of births prior to the present pregnancy (live and stillbirths); for Finland it is the >=1 documented birth in the 5- year LBP
**BRCA1/BRCA2 gene [insertion, deletion, amplification, alteration, fusion, mutation] ***Available from January 1, 2004 onwards in Danish data. For Finland, Staging based on Finnish criteria was available for the whole cohort, TNM data available for only 21,176 women
[1]Ascertained at the time of diagnosis and for pregnancy we used a 9 mo. Lookback
[2]Schizophrenia, schizotypal and delusional disorders, severe (psychotic level) depression, psychosis
[3]Ischemic heart disease, heart failure; thromboembolic events (deep venous thrombosis, pulmonary embolism, stroke, hypertension, hyperlipidaemia
[4]Available for Portugal 2016-2022
[5]Ascertained during 12 mo. follow-up after diagnosis date
[6]Anastrozole, exemestane, letrozole, tamoxifen, fulvestrant and for premenopausal women HER2 inhibitors: trastuzumab, pertuzumab, trastuzumab emtansine, trastuzumab deruxtecan, trastuzumab duocarmazine, margetuximab, neratinib
[7]Ascertained until end of follow-up
[8]Derived from TMN: Stage 0, Abnormal cells are present but have not spread to nearby tissue. Also called carcinoma in situ, or CIS. CIS is not cancer, but it may become cancer; [Stage: local (I), regional (II/III), distant (IV)]. Cancer is present. The higher the number, the larger the cancer tumour and the more it has spread into nearby tissues; For Finland, The Finnish cancer registry uses its own staging method (https://syoparekisteri.fi/assets/files/2021/07/Description-of-the-materials-ofthe-Finnish-Cancer-Registry.pdf)

#### Supplementary Table 3: Demographic and clinical characteristics of incident ALS[1] patients in participating countries (Denmark and Finland, 2000–2021; Portugal, 2005–2022)

| Characteristic | Denmark | | Finland | | Portugal | |
|---|---|---|---|---|---|---|
| | NATIVE | OMOP | NATIVE | OMOP | NATIVE | OMOP |
| | N=2810 | N=2991 | N=5403 | N=5403 | N=4061 | N=4071 |
| Women | 1265 (45%) | 1340 (45%) | 2614 (48%) | 2614 (48%) | 1825 (45%) | 1827 (45%) |
| Men | 1545 (55%) | 1651 (55%) | 2789 (52%) | 2789 (52%) | 2236 (55%) | 2244 (55%) |
| **Age, years** [2] | | | | | | |
| 18 - 34 | 25 (1%) | 34 (1%) | 62 (1%) | 62 (1%) | 58 (1%) | 58 (1%) |
| 35 - 44 | 95 (3%) | 102 (3%) | 147 (3%) | 147 (3%) | 154 (4%) | 154 (4%) |
| 45 - 54 | 285 (10%) | 309 (10%) | 606 (11%) | 606 (11%) | 385 (9%) | 387 (10%) |

| | | | | | | |
|---|---|---|---|---|---|---|
| 55 - 64 | 680 (24%) | 713 (24%) | 1439 (27%) | 1439 (27%) | 907 (22%) | 908 (22%) |
| 65 - 74 | 1060 (38%) | 1131 (38%) | 1834 (34%) | 1834 (34%) | 1435 (35%) | 1438 (35%) |
| >=75 | 655 (23%) | 702 (23%) | 1315 (24%) | 1315 (24%) | 1122 (28%) | 1126 (28%) |
| Median (quartiles) | 68 (60-75) | 68 (59-74) | 67 (59-74) | 67 (59-74) | 69 (61-76) | 68 (60-75) |
| **Year of diagnosis** | | | | | | |
| DK,FL: 2000 - 2010/PT: 2005-2013 | 940 (33%) | 1003 (34%) | 2387 (44%) | 2387 (44%) | 1627 (40%) | 1631 (40%) |
| DK,FL: 2011 - 2021/PT: 2014 - 2022 | 1870 (67%) | 1988 (66%) | 3016 (56%) | 3016 (56%) | 2434 (60%) | 2440 (60%) |
| **Comorbidities** [3] | | | | | | |
| Psychiatric disorders [4] | 15 (1%) | 26 (1%) | 174 (3%) | 174 (3%) | 24 (1%) | 24 (1%) |
| Dementia | 115 (4%) | 157 (5%) | 325 (6%) | 325 (6%) | 249 (6%) | 249 (6%) |
| Cardiovascular diseases [5] | 455 (16%) | 456 (15%) | 1916 (36%) | 1916 (36%) | 1290 (32%) | 1292 (32%) |
| Respiratory diseases | 205 (7%) | 203 (7%) | 656 (12%) | 656 (12%) | 363 (9%) | 364 (9%) |
| Other muscular atrophy types [6] | 20 (1%) | 53 (2%) | NA | NA | 17 (<1%) | 17 (<1%) |
| **Characteristics in 5 years before ALS diagnosis** [3] | | | | | | |
| Fasciculations, cramps, muscle twitching | 45 (2%) | 62 (2%) | 206 (4%) | 185 (3%) | 16 (0%) | 16 (0%) |
| Dysarthris | 95 (3%) | 119 (4%) | 723 (13%) | 719 (13%) | 49 (1%) | 48 (1%) |
| Fronto-temporal dementia | 10 (<1%) | <5 (<1%) | 54 (1%) | 52 (1%) | 13 (0%) | 13 (<1%) |
| Mood disorders (incl. depression, bipolar) | 85 (3%) | 128 (4%) | 273 (5%) | 233 (4%) | 169 (4%) | 174 (4%) |
| Other mild motor impairment | 505 (18%) | 686 (23%) | 1364 (25%) | 1355 (25%) | 33 (1%) | 33 (1%) |
| Absent or pathological brisk reflexes | 0 (0%) | 0 (0%) | 10 (<1%) | 10 (<1%) | NA | 0 (0%) |
| Anxiety | 20 (1%) | 34 (1%) | 84 (2%) | 70 (1%) | 43 (1%) | 43 (1%) |
| **Medication use in 5 years before ALS diagnosis** | | | | | | |
| Riluzole | 1180 (42%) | 1279 (43%) | 311 (6%) | 311 (6%) | NA | NA |
| Antidepressants/mood disorder medication | 1055 (38%) | 1123 (38%) | 1494 (28%) | 1497 (28%) | 399 (75%) | 401 (75%) |
| Benzodiazepines and related drugs | 1050 (37%) | 1185 (40%) | 1569 (29%) | 1572 (29%) | 345 (65%) | 346 (65%) |
| Antiepileptics [incl. gabapentinoids, levetiracetam] | 230 (8%) | 82 (3%) | 520 (10%) | 520 (10%) | 143 (27%) | 105 (20%) |

| | | | | | | |
|---|---|---|---|---|---|---|
| Skeletal muscle relaxants | 445 (16%) | 486 (16%) | 1166 (22%) | 1166 (22%) | 197 (37%) | 198 (37%) |
| Antibiotics | 2125 (76%) | 2295 (77%) | 3731 (69%) | 3731 (69%) | 429 (80%) | 431 (81%) |
| Triamcinolone (+/- lidocaine) | 10 (<1%) | 12 (1%) | 3 (<1%) | 3 (<1%) | NA | NA |
| Hyoscine / Scopolamine / Buscopan | 25 (1%) | 31 (1%) | 14 (<1%) | 14 (<1%) | 13 (2%) | 13 (2%) |
| Cannabis | 0 (0%) | <5 | 0 | 0 | NA | NA |
| **Signals of disease treatment** [7] | | | | | | |
| Intensive care unit service admission with mechanical ventilation | 255 (9%) | 348 (12%) | NA | NA | 76 (2%) | 76 (2%) |
| Non-invasive ventilation | 560 (20%) | 413 (14%) | NA | NA | 880 (22%) | 883 (22%) |
| Riluzole | 1635 (58%) | 1718 (57%) | 1616 (30%) | 1616 (30%) | NA | NA |
| Antiepileptics [incl. gabapentinoids, levetiracetam] | 105 (4%) | 117 (4%) | 348 (6%) | 348 (6%) | 226 (12%) | 149 (8%) |
| Skeletal muscle relaxants | 485 (17%) | 528 (18%) | 492 (9%) | 492 (9%) | 274 (14%) | 306 (16%) |
| Benzodiazepines and related drugs | 1180 (42%) | 1352 (45%) | 1465 (27%) | 1470 (27%) | 634 (33%) | 695 (36%) |
| Hyoscine, Scopolamine, Buscopan | 90 (3%) | 108 (4%) | 12 (<1%) | 12 (<1%) | 33 (2%) | 33 (2%) |
| Triamcinolone +- lidocaine | 0 (0%) | <5 (<1%) | 0 (0%) | 0 (0%) | NA | NA |
| Antidepressants/mood disorder medication | 1200 (43%) | 1284 (43%) | 1841 (34%) | 1843 (34%) | 1018 (52%) | 1117 (57%) |
| **Signals of disease progression** [8] | | | | | | |
| Respiratory infection [pneumonia] | 750 (27%) | 758 (25%) | 1379 (26%) | 1585 (29%) | 1232 (30%) | 786 (19%) |
| Intensive care unit service admission with mechanical ventilation | 440 (16%) | 570 (19%) | NA | NA | 125 (3%) | 120 (3%) |
| Non-invasive ventilation | 805 (29%) | 603 (20%) | NA | NA | 1229 (30%) | 1231 (30%) |
| PEGs (percutaneous gastrostomy) | 380 (13%) | 218 (7%) | 573 (11%) | 608 (11%) | 547 (13%) | 352 (9%) |
| Cachexia (wasting syndrome) | 80 (3%) | 81 (3%) | 10 (<1%) | 10 (<1%) | 162 (4%) | 95 (2%) |
| Acute respiratory failure | 180 (6%) | 188 (6%) | 138 (3%) | 135 (3%) | 682 (17%) | 201 (5%) |
| ALS specific mortality | 2010 (72%) | 2563 (86%) | 3812 (71%) | 3812 (71%) | 1397 (65%) | 1406 (58%) |
| All-cause mortality | 2350 (84%) | 2196 (73%) | 4429 (82%) | 4425 (82%) | 3368 (83%) | 3378 (83%) |

Counts are masked according to each country's privacy-preserving rules

Abbreviations: NA not available

[1] In Denmark can be defined: Progressive spinal paralysis; Progressive bulbar palsy (Bulbar ALS); ALS in the total population was identified : In Denmark with codes: G12.2E, G12.2F, G12.2G, in Finland with : G12.2*, in Portugal with ICD10-CM G1221 and ICD-9-CM 33520

[2]Acertained at the time of diagnosis

[3]Ascertained at time of diagnosis minus 5 years for all data nodes; Finland used diagnoses from specialized healthcare visits collected from the time of diagnosis to minus 5 years and also diagnoses from the Special Reimbursement register from the time of diagnosis to the start of the register (1964) as they are typically recorded only once in the register.
[4]Schizophrenia, schizotypal and delusional disorders, severe (psychotic level) depression, psychosis
[5]Ischemic heart disease, heart failure; thromboembolic events (deep venous thrombosis, pulmonary embolism, stroke, hypertension, hyperlipidaemia)
[6]Including: Progressive muscular atrophy of the spinal type; Progressive muscular atrophy of the myelopathic type; Duchenne-Aran muscular atrophy; Progressive muscular atrophy (PMA)
[7]Ascertained during 12 mo. follow-up after diagnosis date
[8]Ascertained until end of follow-up

**Supplemntary Table 4: Data quality assessment of OMOP-transformed ALS subset from the Danish registries data.**

| | **Verification** | | | | **Validation** | | | | **Total** | | | |
|---|---|---|---|---|---|---|---|---|---|---|---|---|
| | Pass | Fail | Total | % Pass | Pass | Fail | Total | % Pass | Pass | Fail | Total | % Pass |
| Plausibility | 507 | 8 | 515 | 98% | 291 | 0 | 291 | 100% | 798 | 8 | 806 | 99% |
| Conformance | 747 | 60 | 807 | 93% | 116 | 3 | 119 | 97% | 863 | 63 | 926 | 93% |
| Completeness | 402 | 48 | 450 | 89% | 18 | 1 | 19 | 95% | 420 | 49 | 469 | 90% |
| Total | 1656 | 116 | 1772 | 93% | 425 | 4 | 429 | 99% | 2081 | 120 | 2201 | 95% |

**Supplementary Table 5: Data quality assessment of OMOP-transformed BC subset from the Danish registries data.**

| | **Verification** | | | | **Validation** | | | | **Total** | | | |
|---|---|---|---|---|---|---|---|---|---|---|---|---|
| | Pass | Fail | Total | % Pass | Pass | Fail | Total | % Pass | Pass | Fail | Total | % Pass |
| Plausibility | 507 | 8 | 515 | 98% | 288 | 3 | 291 | 99% | 795 | 11 | 806 | 99% |
| Conformance | 744 | 63 | 807 | 92% | 115 | 4 | 119 | 97% | 859 | 67 | 926 | 93% |
| Completeness | 401 | 49 | 450 | 89% | 18 | 1 | 19 | 95% | 419 | 50 | 469 | 89% |
| Total | 1652 | 120 | 1772 | 93% | 421 | 8 | 429 | 98% | 2073 | 128 | 2201 | 94% |

**Supplementary Table 6: Data quality assessment of OMOP-transformed ALS subset from the Portuguese registries data.**

| | Verification | | | | Validation | | | | Total | | | |
|---|---|---|---|---|---|---|---|---|---|---|---|---|
| | Pass | Fail | Total | % Pass | Pass | Fail | Total | % Pass | Pass | Fail | Total | % Pass |
| Plausibility | 513 | 2 | 515 | 100% | 277 | 14 | 291 | 95% | 790 | 16 | 806 | 98% |
| Conformance | 784 | 54 | 812 | 97% | 118 | 1 | 119 | 99% | 902 | 55 | 931 | 97% |
| Completeness | 440 | 45 | 449 | 98% | 14 | 5 | 19 | 74% | 454 | 50 | 468 | 97% |
| Total | 1737 | 101 | 1776 | 98% | 409 | 20 | 429 | 95% | 2146 | 121 | 2205 | 97% |

**Supplementary Table 7: Data quality assessment of OMOP-transformed BC subset from the Portuguese registries data.**

| | Verification | | | | Validation | | | | Total | | | |
|---|---|---|---|---|---|---|---|---|---|---|---|---|
| | Pass | Fail | Total | % Pass | Pass | Fail | Total | % Pass | Pass | Fail | Total | % Pass |
| Plausibility | 515 | 0 | 515 | 100% | 281 | 10 | 291 | 97% | 796 | 10 | 806 | 99% |
| Conformance | 776 | 62 | 812 | 96% | 118 | 1 | 119 | 99% | 894 | 63 | 931 | 96% |
| Completeness | 442 | 43 | 449 | 98% | 15 | 4 | 19 | 79% | 457 | 47 | 468 | 98% |
| Total | 1733 | 105 | 1776 | 98% | 414 | 15 | 429 | 97% | 2147 | 120 | 2205 | 97% |

**Supplementary Table 8: Data quality assessment of OMOP-transformed ALS subset from the Finnish registries data.**

| | Verification | | | | Validation | | | | Total | | | |
|---|---|---|---|---|---|---|---|---|---|---|---|---|
| | Pass | Fail | Total | % Pass | Pass | Fail | Total | % Pass | Pas s | Fail | Total | % Pass |
| Plausibility | 505 | 10 | 515 | 98% | 288 | 3 | 291 | 99% | 793 | 13 | 806 | 98% |
| Conformance | 747 | 60 | 807 | 93% | 119 | 0 | 119 | 100% | 866 | 60 | 926 | 94% |
| Completeness | 405 | 45 | 450 | 90% | 12 | 7 | 19 | 63% | 417 | 52 | 469 | 89% |
| Total | 1657 | 115 | 177 2 | 94% | 419 | 1 0 | 429 | 98% | 2076 | 125 | 220 1 | 94% |

**Supplementary Table 9: Data quality assessment of OMOP-transformed BC subset from the Finnish registries data.**

| | Verification | | | | Validation | | | | Total | | | |
|---|---|---|---|---|---|---|---|---|---|---|---|---|
| | Pass | Fail | Total | % Pass | Pass | Fail | Total | % Pass | Pass | Fail | Total | % Pass |
| Plausibility | 505 | 10 | 515 | 98% | 277 | 14 | 291 | 95% | 782 | 24 | 806 | 97% |

| | | | | | | | | | | | | |
|---|---|---|---|---|---|---|---|---|---|---|---|---|
| Conformance | 741 | 66 | 807 | 92% | 119 | 0 | 119 | 100% | 860 | 66 | 926 | 93% |
| Completeness | 403 | 47 | 450 | 90% | 11 | 8 | 19 | 58% | 414 | 55 | 469 | 88% |
| Total | 1649 | 123 | 1772 | 93% | 407 | 22 | 429 | 95% | 2056 | 145 | 2201 | 93% |
